# Physiology of cardiomyocyte injury in COVID-19

**DOI:** 10.1101/2020.11.10.20229294

**Authors:** Mustafa M. Siddiq, Angel T. Chan, Lisa Miorin, Arjun S. Yadaw, Kristin G. Beaumont, Thomas Kehrer, Kris M. White, Anastasija Cupic, Rosa E. Tolentino, Bin Hu, Alan D. Stern, Iman Tavassoly, Jens Hansen, Pedro Martinez, Nicole Dubois, Christoph Schaniel, Rupa Iyengar-Kapuganti, Nina Kukar, Gennaro Giustino, Karan Sud, Sharon Nirenberg, Patricia Kovatch, Joseph Goldfarb, Lori Croft, Maryann A. McLaughlin, Edgar Argulian, Stamatios Lerakis, Jagat Narula, Adolfo García-Sastre, Ravi Iyengar

## Abstract

COVID-19 affects multiple organs. Clinical data from the Mount Sinai Health System shows that substantial numbers of COVID-19 patients without prior heart disease develop cardiac dysfunction. How COVID-19 patients develop cardiac disease is not known. We integrate cell biological and physiological analyses of human cardiomyocytes differentiated from human induced pluripotent stem cells (hiPSCs) infected with SARS-CoV-2 in the presence of interleukins, with clinical findings, to investigate plausible mechanisms of cardiac disease in COVID-19 patients. We infected hiPSC-derived cardiomyocytes, from healthy human subjects, with SARS-CoV-2 in the absence and presence of interleukins. We find that interleukin treatment and infection results in disorganization of myofibrils, extracellular release of troponin-I, and reduced and erratic beating. Although interleukins do not increase the extent, they increase the severity of viral infection of cardiomyocytes resulting in cessation of beating. Clinical data from hospitalized patients from the Mount Sinai Health system show that a significant portion of COVID-19 patients without prior history of heart disease, have elevated troponin and interleukin levels. A substantial subset of these patients showed reduced left ventricular function by echocardiography. Our laboratory observations, combined with the clinical data, indicate that direct effects on cardiomyocytes by interleukins and SARS-CoV-2 infection can underlie the heart disease in COVID-19 patients.

**One Sentence Summary:** Cardiomyocytes derived from human induced pluripotent stem cells treated with interleukins and infected with SARS- CoV- 2 in cultures, show increased release of troponin, disorganization of myofibrils, and changes in beating mirroring specific pathologies in some COVID-19 patients.

## Introduction

SARS-CoV-2 infections have created a global COVID-19 pandemic that continues unabated. With over 50 million cases worldwide (1), COVID-19 has affected people in every country. This is in contrast to other coronavirus outbreaks such as SARS (2) and MERS (3) that were geographically limited. Significant numbers of patients with COVID-19 display multi-organ pathophysiology. There is substantial evidence that a significant proportion of COVID-19 patients develop both cardiac (4-6) and kidney dysfunction (7, 8). Such multi-organ pathophysiology is perhaps not entirely surprising since the receptor for SARS-CoV-2 is the angiotensin-converting enzyme 2 (ACE2) protein (9) that is widely expressed in many tissues including the heart and kidney (10)

Myocardial injury associated with COVID-19 has been observed in substantial numbers of patients (11). Biomarkers of injury include elevated troponin and BNP protein levels in blood (12) and abnormal echocardiography (13, 14). The reasons for the myocardial injury remain unclear, and autopsy results have shown that COVID-19 patients show sparse infection of cardiac tissue and do not show the typical hallmarks of viral myocarditis (15). Myocardial injury can have multiple causes including formation of microthrombi that have been observed in COVID-19 patients (16) and oxygen insufficiency due to pulmonary disease that is commonly observed, and in our analyses is an important predictor of mortality (17). Another feature observed in the clinical data is that most COVID-19 patients have elevated interleukins (18). Interleukins are known to affect cardiomyocyte function (19). Multiple reports have indicated that human cardiomyocytes can be infected by SARS-CoV-2 (20-22). In this study, we integrated findings from analyses of fully anonymized clinical data on COVID-19 patients from the Mount Sinai Health System, with those from studies on hiPSC-derived ventricular cardiomyocytes in culture, infected with SARS–CoV-2 in the absence and presence of interleukins. We demonstrate that changes in cardiomyocyte cell biology, such as myofibril organization, and in physiology, such as release of troponin and erratic beating of cardiomyocytes, can account for key features of myocardial injury observed in patients. These observations demonstrate that direct injury by virus to cardiac tissue during COVID -19 can play a role in decreased cardiac function.

## Results

### COVID-19 patients without prior heart disease show elevated troponin levels

During the early phase of the pandemic in New York City in spring 2020, substantial numbers of patients were over 55 years old (23). This was true of patients in the Mount Sinai Health System as well (Supplementary Table 1). Hence, we parsed the deidentified clinical data compiled by the Mount Sinai Data Warehouse (MSDW) to identify COVID-19 patients with and without prior history of heart disease. For this, we used three co-morbidities coronary artery disease, atrial fibrillation and heart failure listed in the MSDW data for each patient as prior indications of heart disease. We parsed the COVID -19 patients to determine the size of the subset that did not have prior heart disease but show cardiac dysfunction during COVID-19. We used elevated troponin I levels in blood as a measure of cardiac dysfunction (24). 32.25% (1020/3163) show elevated blood troponin levels (Figure 1A). Since elevated blood troponin levels can occur due to kidney dysfunction, patients were binned with estimated glomerular filtration rate (eGFR) less than and greater than or equal to 30 ml/min. 21% (281/1341) of the patients with eGFR ≥30 ml/min show elevated levels of blood troponin (Figure 1B right panel). Some of these 281 patients had comorbities that are indicated in the right panel in Figure 1B The overall numbers indicate 44% of patients (901/2562) of COVID-19 patients who have recorded eGFR values in MSDW have diminished kidney function (eGFR <30). Comorbidities for some of the patients at different levels of troponin are shown in Fig 1B right panel. Overall, we find a significant number of COVID-19 patients with normal kidney function show elevated blood troponin I levels. In order to further verify that blood troponin I levels, represent cardiac dysfunction we sought to determine, in an unbiased manner, other clinical features associated with elevated troponin I in COVID-19 patients. For this we used machine-learning models to predict the top ranked features in patients with elevated troponin I levels. We had previously used this approach to predict clinical indicators of probable mortality in COVID-19 patients (17). The flow chart Figure 1C-I shows how we developed and used the machine-learning model. The XGBoost algorithm model gave good predictive performance with an AUC (area under the curve) of 0.846 (Figure 1C-II). Details of model comparison and performance are shown in Supplementary figures 1, 2 and 3 and Supplementary Tables 1 and 2. We used the XGBoost model to identify the top ranked features that predict elevated blood troponin. The list is shown in Figure 1C-III. It can be seen that three of top five features BNP, Systolic BPmin and heart rate directly relate to cardiac function. We conclude that release of troponin can be used to evaluate cardiomyocyte injury upon infection with SARS-CoV-2.

**Figure 1.**
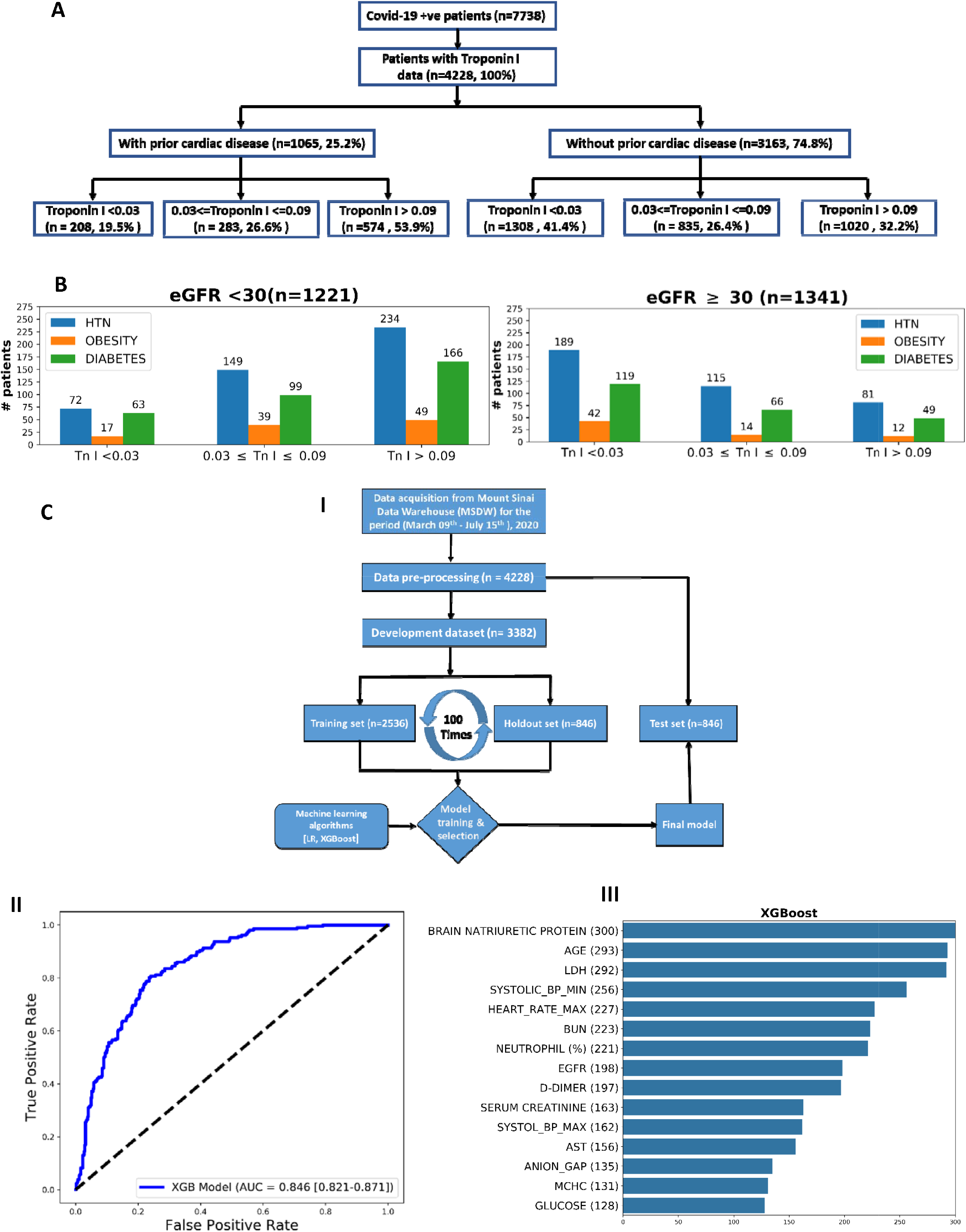
COVID-19 patients without prior history of cardiac disease have elevated troponin I levels, and other clinical characteristics associated with cardiac dysfunction. Clinical data of COVID-19 positive patients (with encounter data, labs and vital signs, n = 7738) downloaded from Mount Sinai Data Warehouse on July 15^th^, 2020. **(A)** Flow chart: COVID-19 patients for whom Troponin I measurements were available (n= 4228) were further divided into patients with and without prior cardiac disease. Patients having one or more of following three comorbidities or prior histry: 1. CORONARY ARTERY DISEASE, 2. ATRIAL FIBRILLATION 3. HEART FAILURE were binned as patients as “with prior cardiac disease”. Each category was further divided into subgroups with respect to troponin levels using standard clinical cutoffs. 32.2% (1020/3163) of COVID-19 patients without prior history of heart disease have clinically significant elevated (>0.09 ng/ml) levels of troponin I. **(B)** Patients without prior cardiac disease were classified with respect to kidney function using eGFR values (< or ≥) 30 and binned for comorbidities (Hypertension, Obesity, Diabetes) in the three cohorts with different Troponin I levels. **(C)** We developed predictive machine learning models to identify clinical features that predict elevated levels of troponin I. The workflow for model development is shown in **Figure 1-C-I:** After pre-processing, data for patients with COVID-19 with troponin I data (n=4228) were randomly divided in an 80:20 ratios into a prediction model development dataset (n=3382) and an independent retrospective validation dataset (test dataset; n=846). For prediction model training and selection, the development dataset was further randomly split into a 75% training dataset (n=2536) and a 25% holdout dataset (n=846). We ran imputation model on the training set to obtain an optimum missing value imputation cutoff which is 35% for this model (Suppl. fig S1). We used a recursive feature elimination method to obtain the optimum number of features to reach plateau (Supplementary fig S2). We tested two classification algorithms, and the XGBoost (eXtreme Gradient Boosting) classification algorithm performed better than logistic regression (Supplementary fig S3). The final predictive model was validated on the test dataset; n= 846). **C-II:** Evaluation results for the test set are shown in terms of the ROC curves obtained, as well as their AUC scores, with 95% confidence interval in parentheses. **C-III:** The top fifteen predictive features identified using the recursive feature elimination method for XGBoost classification algorithms across the three independent sets of 100 runs used to select the most discriminative features, and train the corresponding candidate prediction models; the values in parentheses indicate the number of times the feature was selected as top ranked in the development dataset.

**Figure 2.**
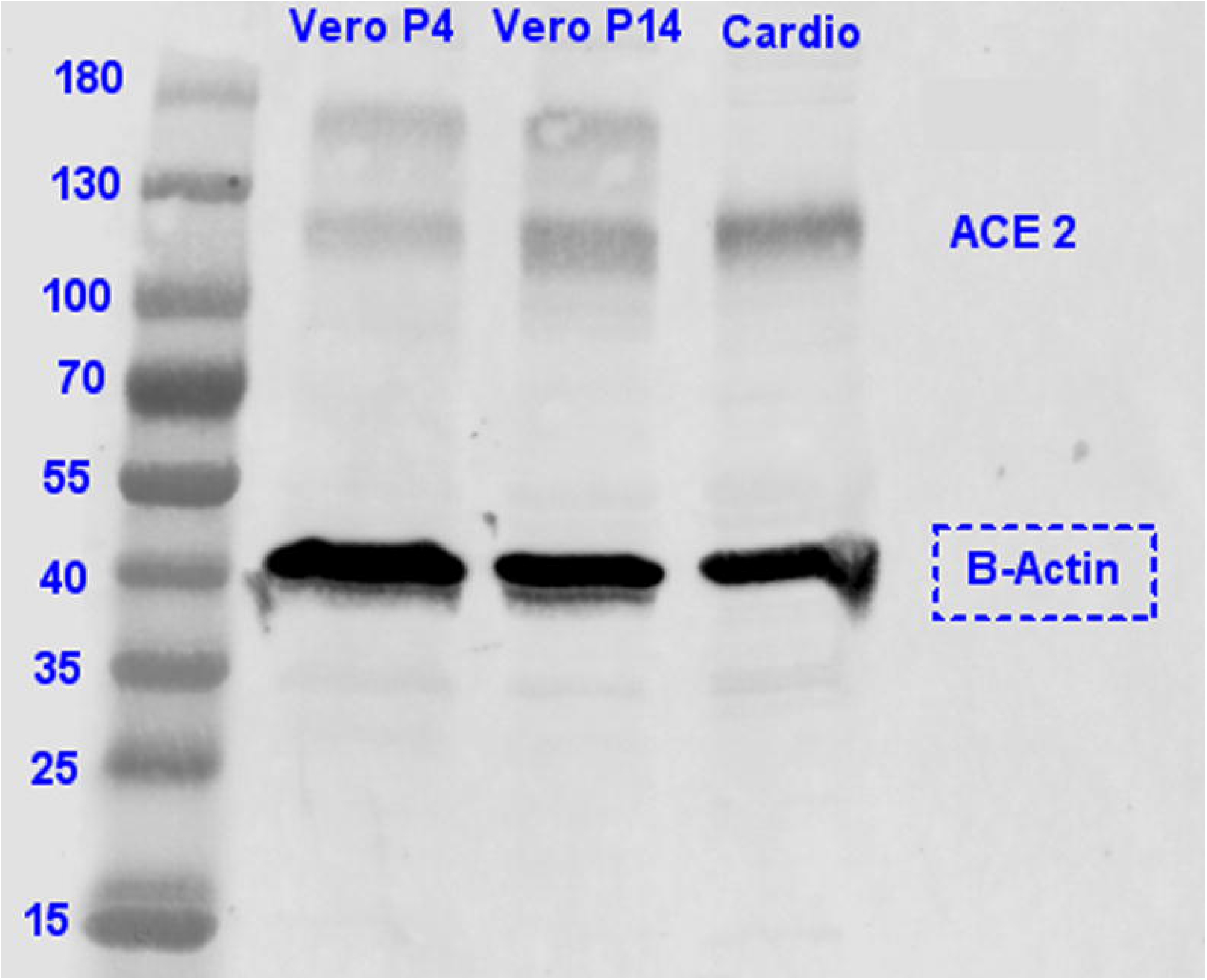
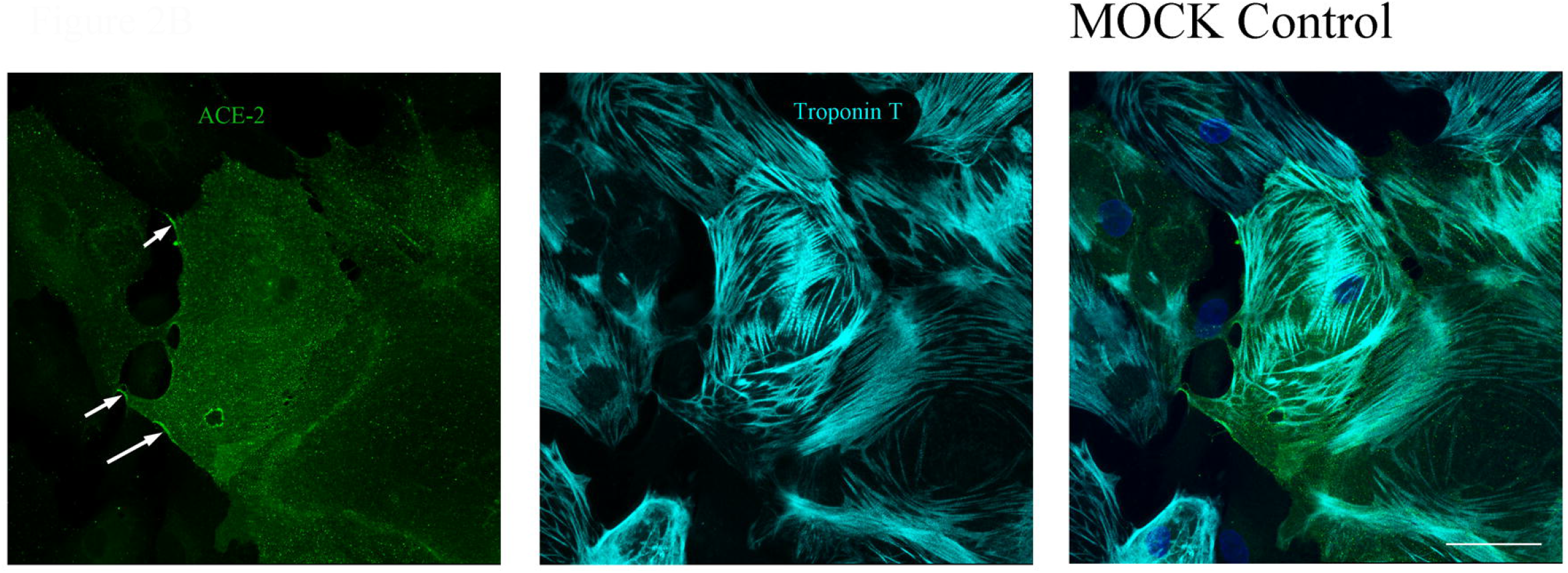
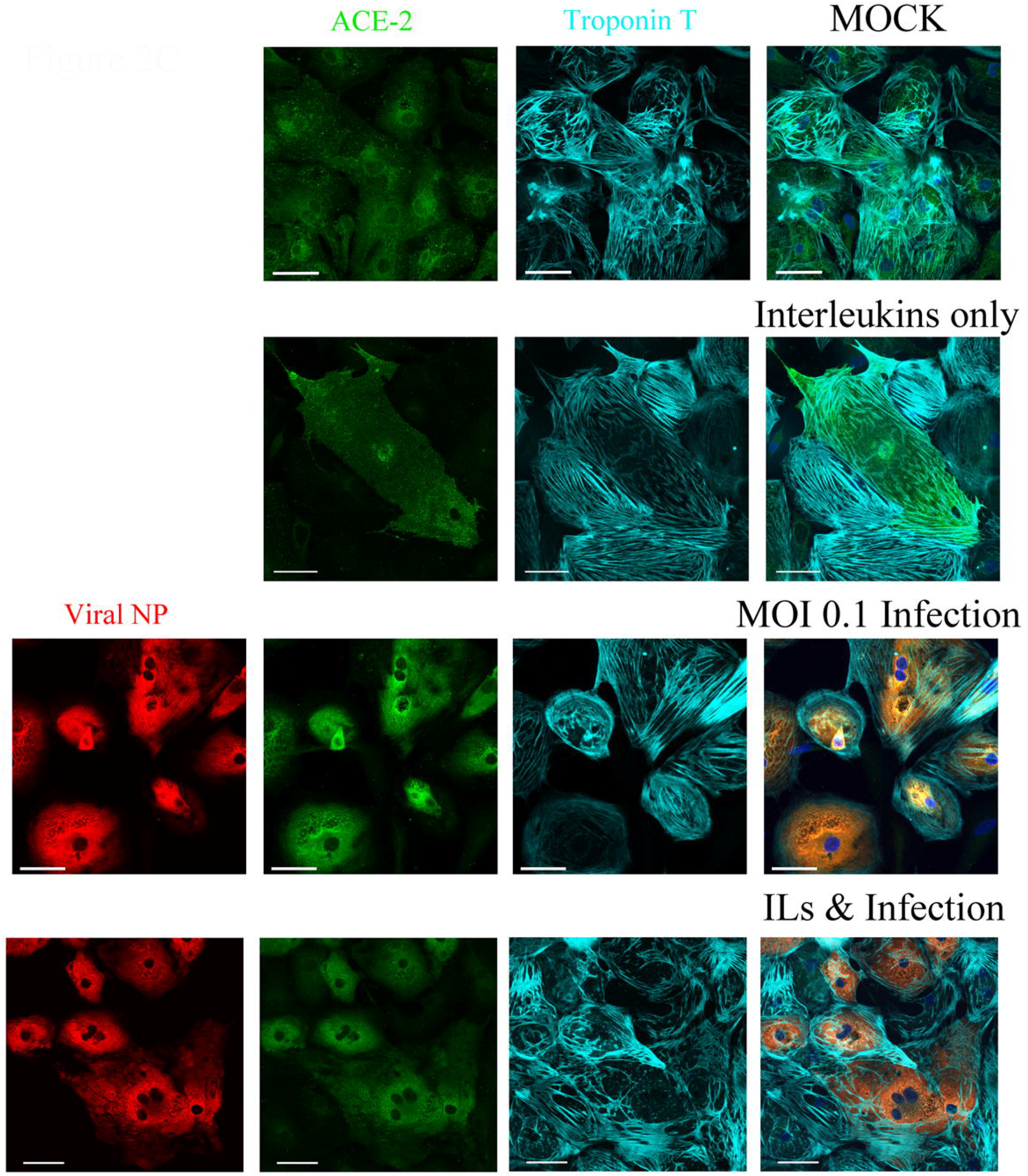
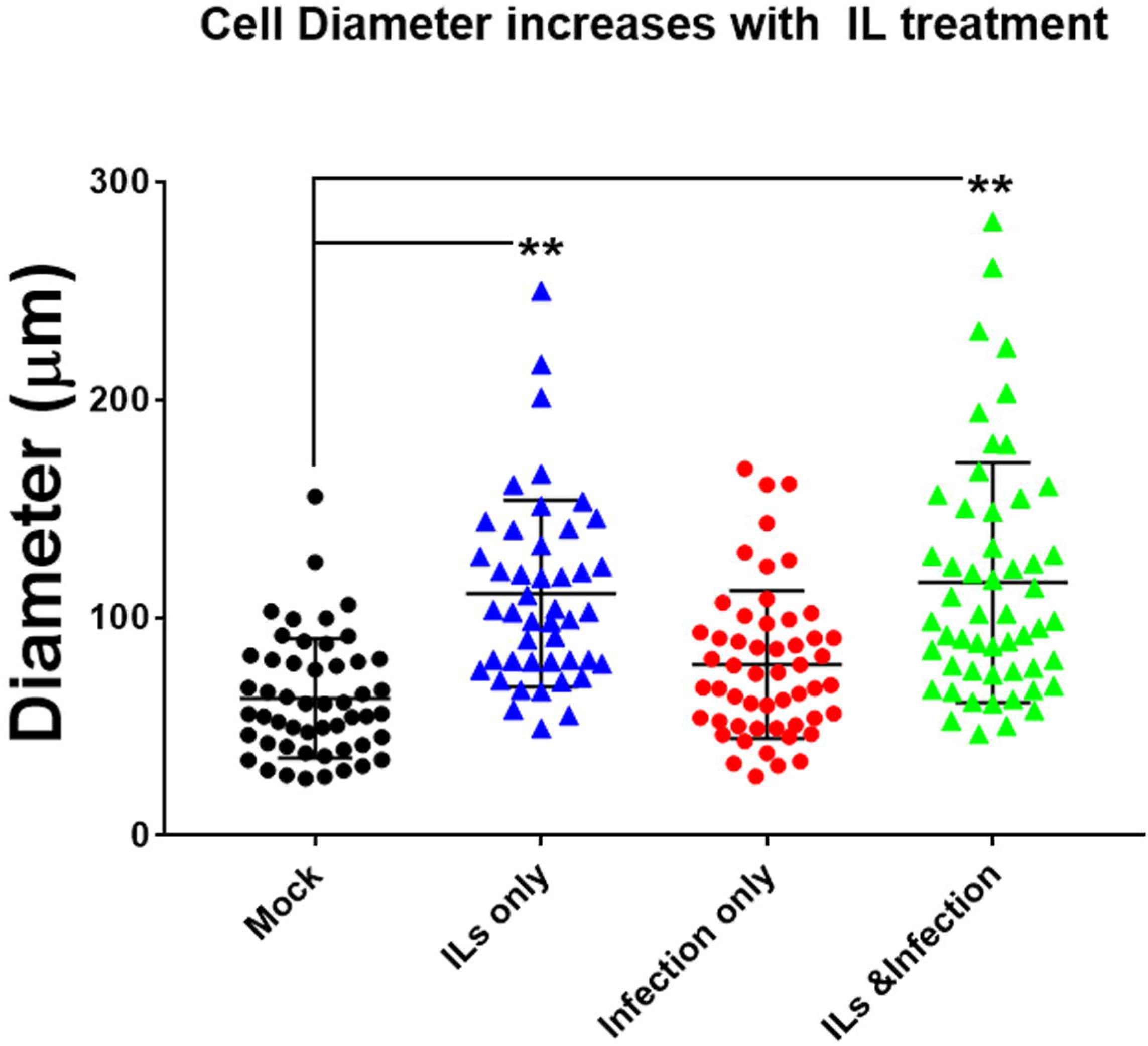
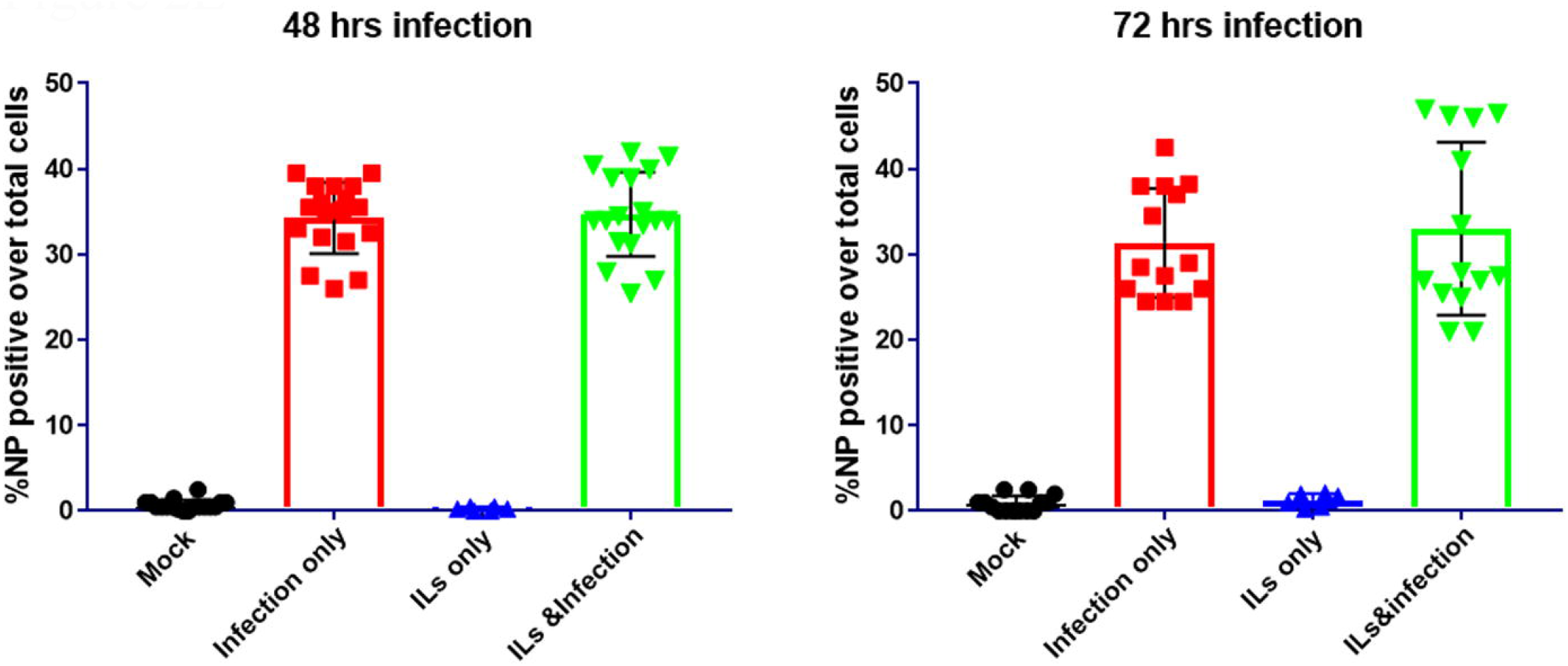
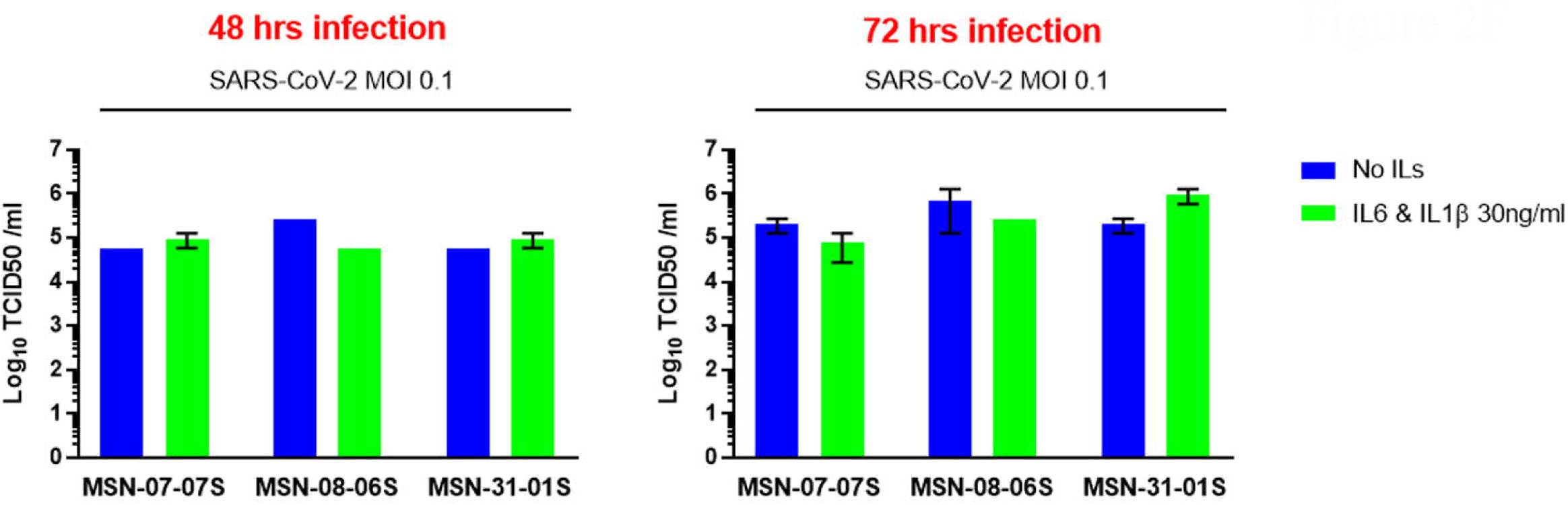
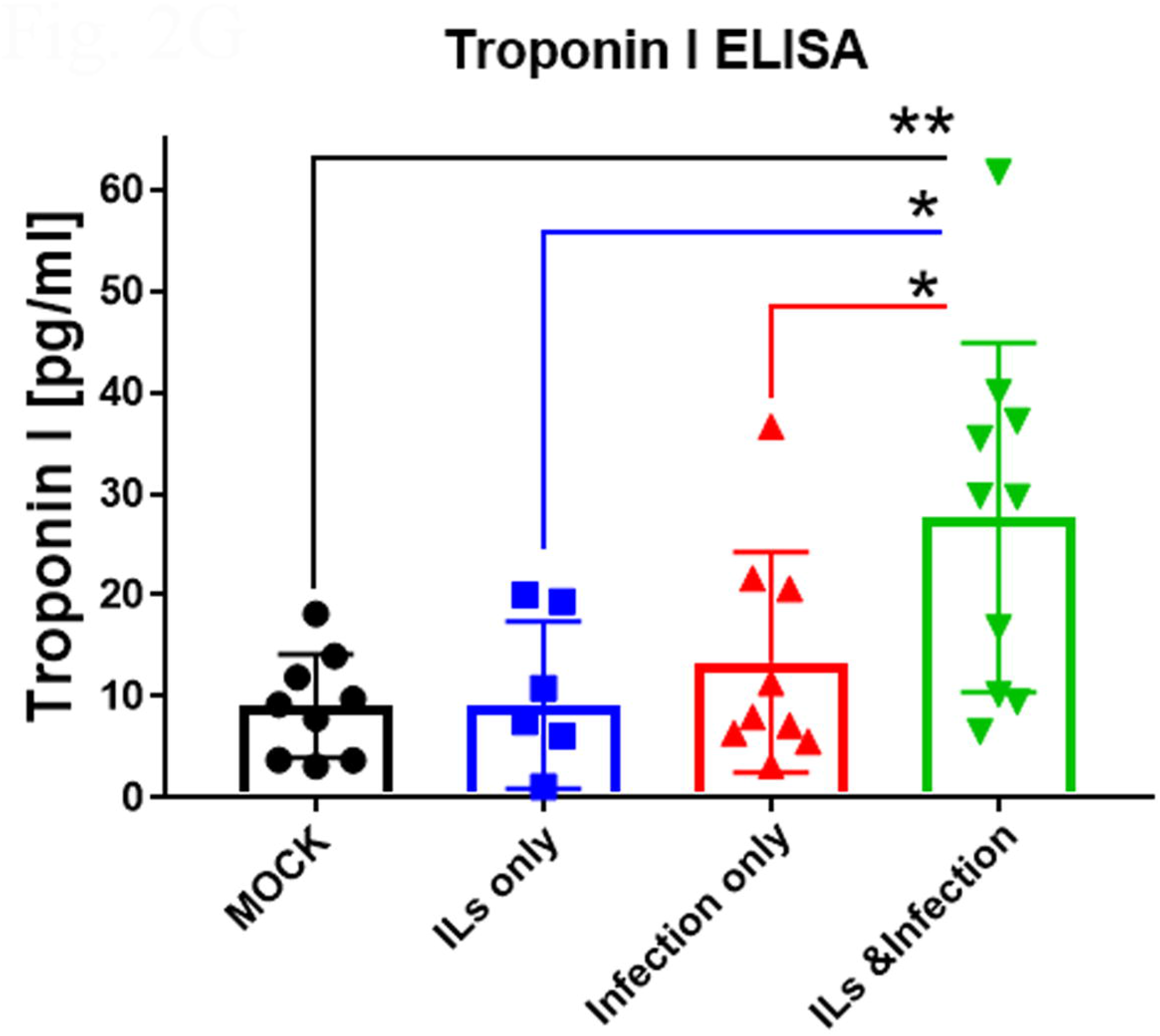
Characterization of hiPSC-derived ventricular cardiomyocytes infected by SARS-CoV-2. **h**iPSC cells reprogrammed from skin fibroblasts of healthy subjects were differentiated into beating ventricular cardiomyocytes (CMs), and after culturing for 30days were used in these experiments. (**A**) Expression of ACE 2 protein. By immunoblotting, the expression of ACE-2 at the predicted size of 120kD was identified in cardiomyocytes (cardio) compared to the standard monkey Vero cell line at two different passages (P4 & P14). (**B**) Immunostaining of CMs for ACE-2 (Green) and Troponin T (Cyan). Although all cells express ACE2, we find only some cells express ACE-2 on the plasma membrane, as indicated by the arrows (left-most panel). The CMs express Troponin T (Cyan) which is seen as ordered myofibrils (middle panel). Overlay of the two-stains is shown in the right panel. The scale bar in the composite image is 50µm. (**C**) Top panels-Mock infected cardiomyocytes express ACE-2 and Troponin T, nuclei are stained by Hoescht. Level 2 panels - CM treated with 30ng/ml each of IL-6 and IL-1β express ACE-2 and the cells appear larger, and the Troponin T appeared more disorganized. Level 3 panels - CM can be infected with SARS-CoV-2 at a MOI 0.1 as detected by positive immunostaining for viral NP (Red), all infected cells were positive for ACE-2 with some cells having notable amounts of Troponin T disruption. Level -4 panels infected with SARS-CoV-2 in the presence of ILs also show increase in cell diameter and disruption in Troponin T organization. The scale bar in the composite image is for 50µm. (**D**) The changes in cell diameter with ILs and/or infection were measured using ImageJ. Cell diameter for two different CM lines were measured and plotted, We observe a statistically significant increase in cell size with infection but a larger increase in cell diameter with ILs only or ILs and infection with SARS-CoV-2. (**E**) To confirm that CM were being productively infected and shedding SARS-CoV-2 into the culture media, we collected supernatants from 3 different CM lines 48 and 72hrs post-infection, with or without ILs and performed a TCID50 plaque assay. We observed that CM lines are infected and the level of infection, as assessed by virus release in to culture medium did not increase with the addition of IL-6 and IL-1β. (**F**) We counted the numbers of cells stained by N protein antibody as a measure of infectivity. Each point is representative from one well of a 96 well plate with 10,000 CM cells. The bar graph is the average of four different CM lines measuring viral NP protein positive cells over total number of cells. Counting was done in an automated fashion using the InCell Analyzer. ILs do not increase the percentage of CM cells infected by SARS-CoV-2. (**G**) Three different CM lines were treated with ILs and/or infection with SARS-CoV-2 at a MOI of 0.1 for 72hrs. The supernatants were collected and used for the measurement of levels of Troponin I released into the media. We found that ILs or infection with SARS-CoV-2 did not significantly release Troponin I into the media. However, infection in the presence of ILs significantly increased release of Troponin into the culture media. Comparing IL treatment or Infection only to ILs with infection, we see there was significant increase in Troponin release when we combined ILs with infection. Statistics are ANOVA with Bonferroni multiple comparison test, *p<0.05 and **p<0.01.

**Figure 3.**
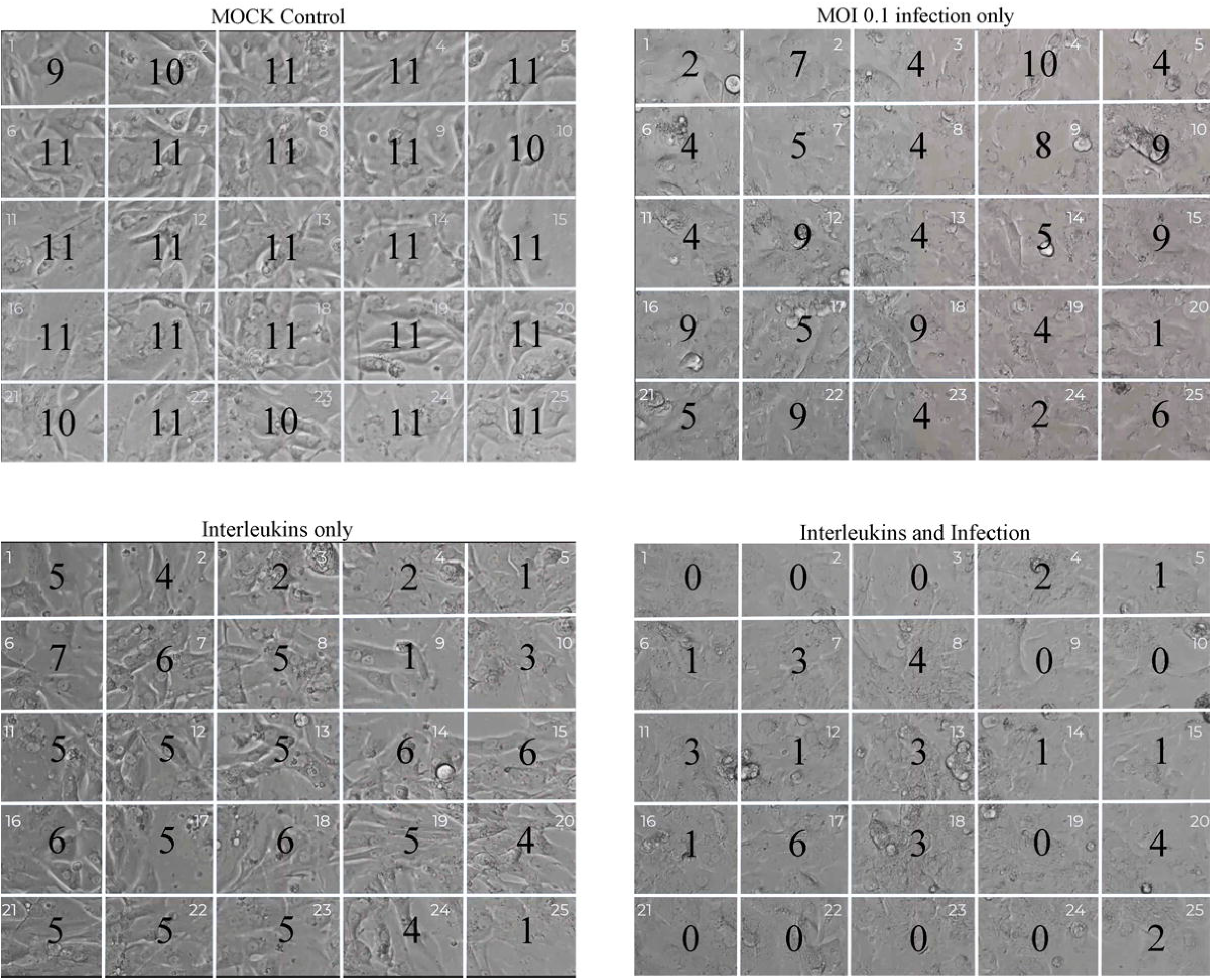
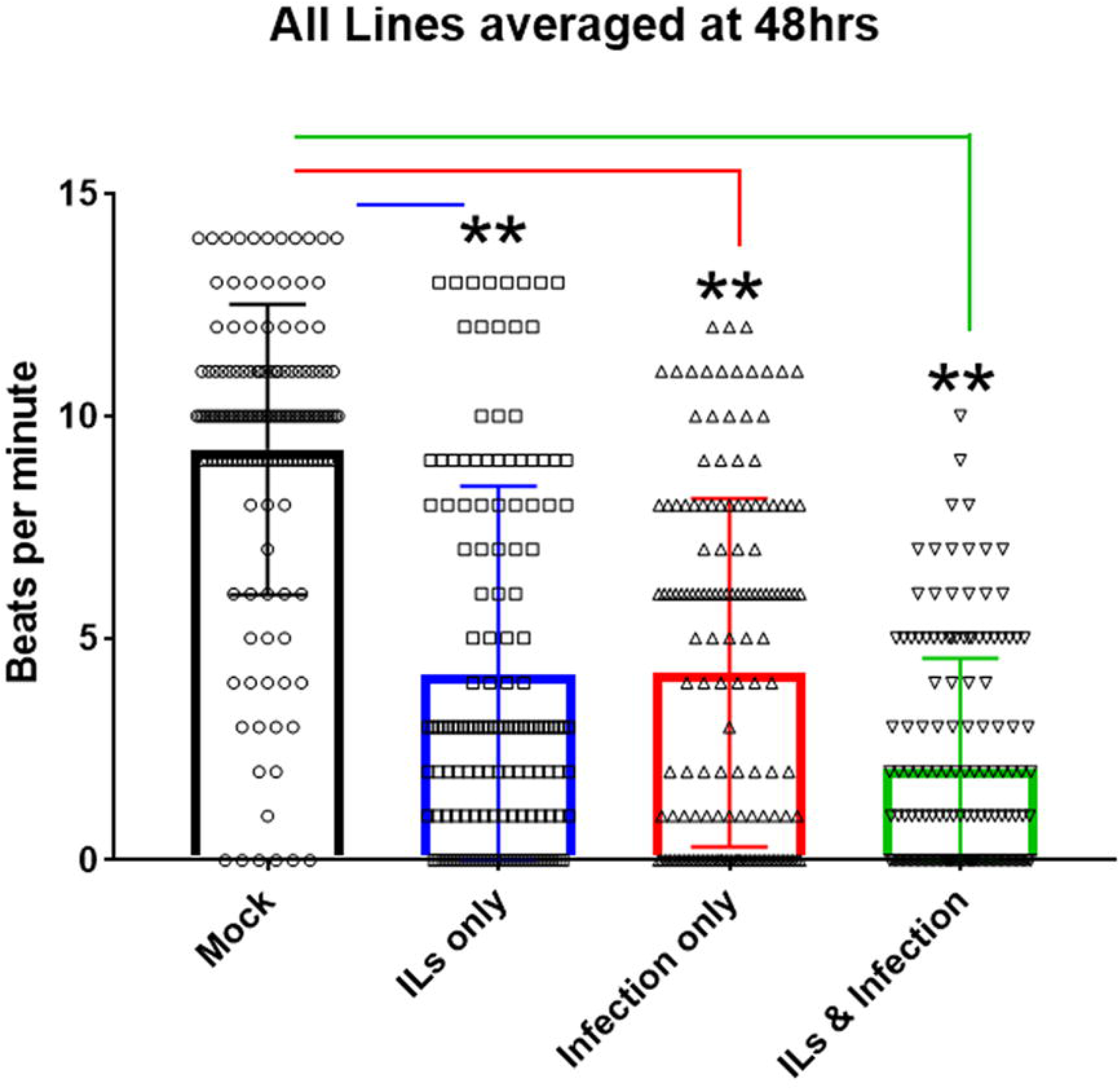
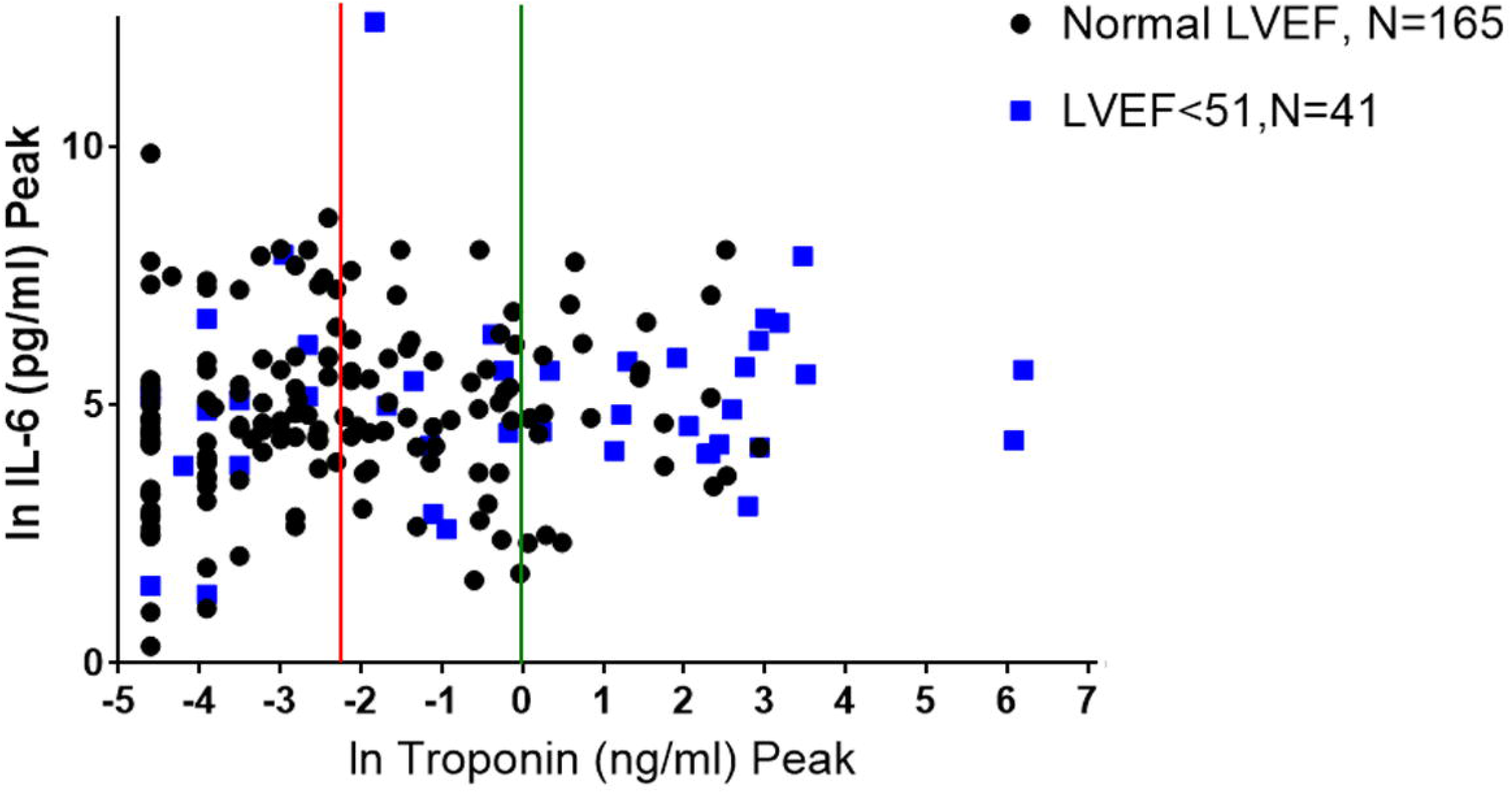
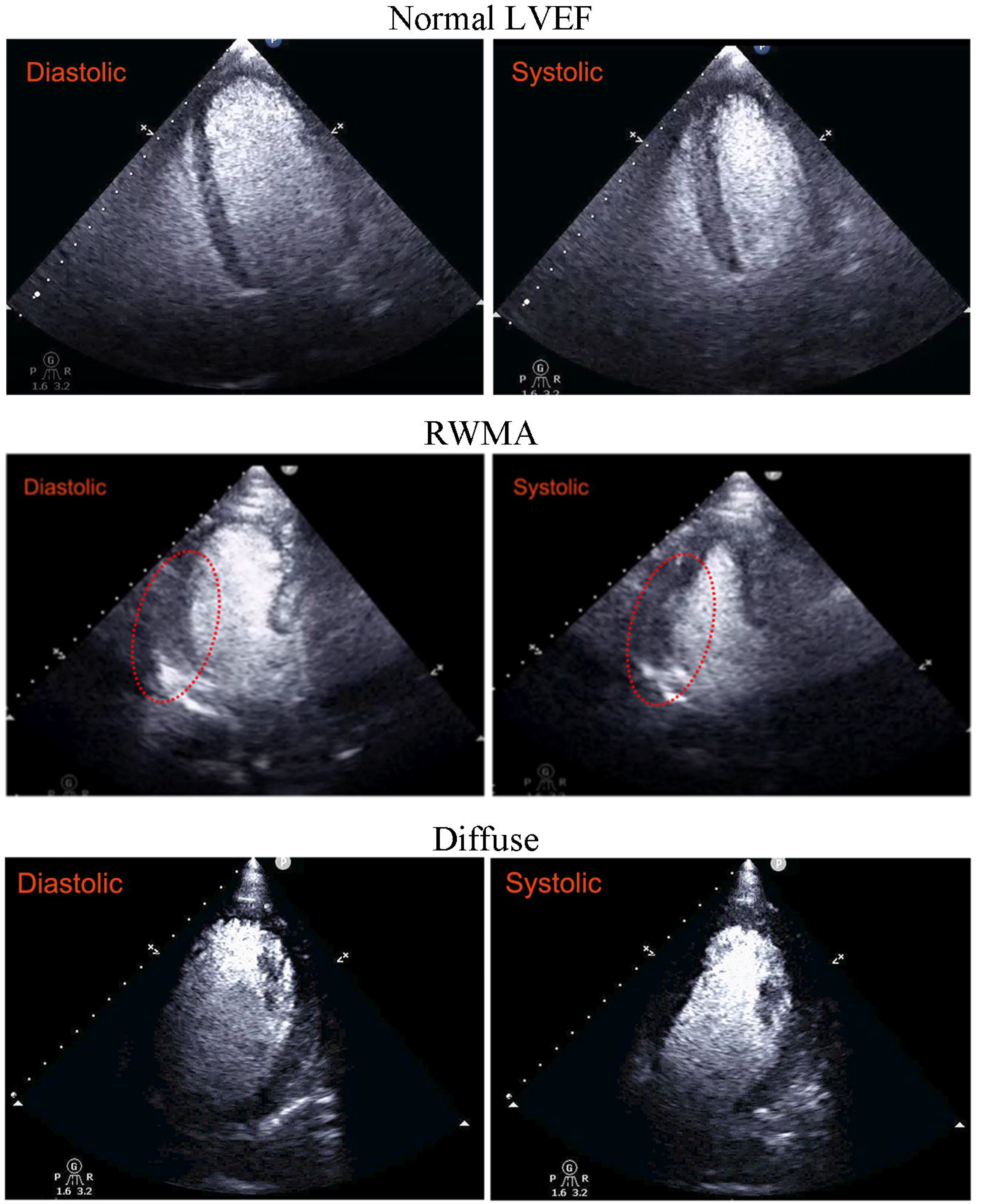
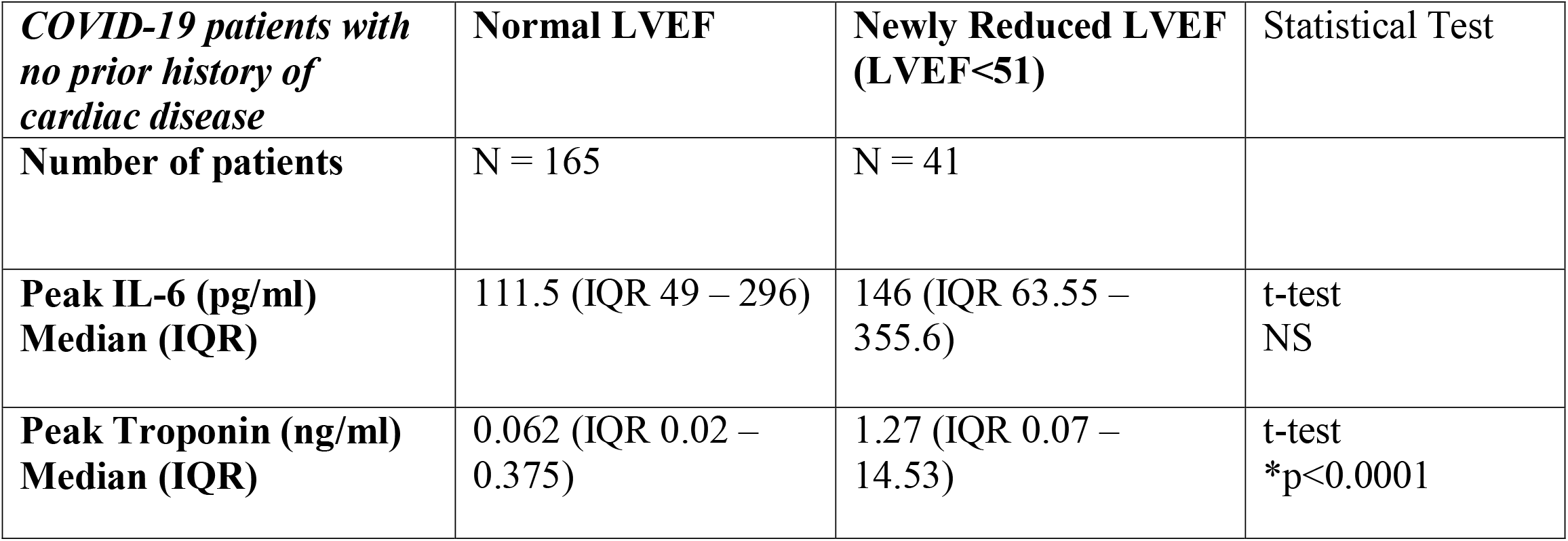
Effect of SARS-CoV-2 infection on beating of ventricular cardiomyocytes in cultures, and cardiac output in COVID-19 patients. (**A**) Three different CM lines were recorded for their beating in culture with ILs and/or infection in a biosafety level-3 facility. These CM lines were in culture for 45-60days. Onto the movies that we recorded, we juxtaposed a 5×5 grid to count the number of beats per min, for each cell line at 48hrs post-infection. The number in each of the 25 grids (beats per minute) is the average of two experiments for one representative CM line, for each condition. We observed that infection alone can slow down the beating, as could ILs treatment. The strongest attenuation of beating was observed with SARS-CoV-2 infection in the presence of ILs. Movie 1 shows the beating of the CMs under various conditions. (**B**) Summary of counts of the beating in three different 3 CM lines are shown Beat measurement in each area of the grid is represented as an open dot and mean and SEM are shown. Asterisks indicate significance at p < 0.001 by ANOVA with Bonferroni multiple comparison test. Though all conditions significantly attenuated beating, the most robust effect was combining ILs with infection with SARS-CoV-2. **C**) COVID-19 patients without prior cardiac disease were divided into two groups, one with normal LVEF (N=165) and those with decreased LVEF<51 (N=41). We plotted the peak Troponin I (ng/ml) using a natural log scale (LN) and peak IL-6 (pg/ml). The red line indicates where Troponin levels are at 0.1 ng/ml (LN = -2.3) and the green line at 1 ng/ml (LN = 0). For the group with LVEF<51, 20 out of 41 (50%) have Troponin levels >1ng/ml, while for those with normal LVEF it is 22/165 (13.3%). Another group of patients with LVEF<51, 6/41 (14.6%), have troponin I levels greater than 20ng/ml (LN = 3); none of the patients with normal LVEF are above 20ng/ml. For troponin I, the difference is very significant by t-test, p<0.0001, when comparing normal LVEF to the LVEF<51 group. **(D-F)** Echocardiogram clips showing Diastolic and Systolic states. <u>(**D**</u>) Normal demonstrates an apical 4-chamber view (after administration of an ultrasonic enhancing agent) from a transthoracic echocardiogram obtained from a COVID-19 patient. The findings are consistent with preserved left ventricular ejection fraction and no regional wall motion abnormalities (diastolic and systolic frames). The end-diastolic and end-systolic volumes are 104ml and 35ml, respectively with a left ventricular ejection fraction of 66%; (<u>**E**</u>) Regional wall motion abnormality (RWMA) demonstrates an apical 3-chamber view (after administration of an ultrasonic enhancing agent) from a transthoracic echocardiogram obtained from a COVID-19 patient. The findings are consistent with basal and mid infero-lateral wall hypokinesis despite preserved left ventricular ejection fraction (diastolic and systolic frames). The RWMA is highlighted by the red oval. The end-diastolic and end-systolic volumes are 102ml and 50ml, respectively with a left ventricular ejection fraction of 51%; (<u>**F**</u>) diffuse demonstrates an apical 4-chamber view (after administration of an ultrasonic enhancing agent) from a transthoracic echocardiogram obtained from a COVID-19 patient. The findings are consistent with diffuse left ventricular wall hypokinesis and mildly decreased left ventricular ejection fraction. The end-diastolic and end-systolic volumes are 160ml and 90ml, respectively with a left ventricular ejection fraction of 44%. (<u>**G**</u>) Summary of cardiac dysfunction effects in COVID-19 patients without prior cardiac disease. Data are from two different hospitals within the Mount Sinai Health System. Data indicate that COVID-19 patients include those with normal left ventricle ejection fraction (LVEF) and with newly reduced LVEF (LVEF<51), and we have the averaged peak of IL-6 and Troponin I for these patients. The t-test for Troponin between the two groups (Normal LVEF and newly reduced LVEF) was significantly different as determined by t-test.

### Cardiomyocytes from hiPSCs when infected with SARS-CoV-2 in the presence of ILs are damaged and release troponin

We have developed a library of hiPSCs from a racially/ethnically diverse population of healthy subjects (25). Since the clinical data analyses in Figure 1 indicate that COVID-19 patients without prior indication of heart disease can develop cardiac dysfunction, we used hiPSCs from volunteers who had been clinically classified as healthy by three physicians using a variety of criteria including electrocardiograms (25) to obtain ventricular cardiomyocytes for the cell biological and physiological analyses. Differentiated ventricular cardiomyocytes from multiple hiPSC lines were used in these studies. In all cases we obtained near homogeneous cardiomyocyte populations by lactate selection (26). In these hiPSC-derived CMs, we first determined the expression of ACE2, the SARS-CoV-2 receptor, by immunoblotting. We compared the CMs to Vero E6 cells, an African green monkey kidney cell line known to express ACE-2 and to support robust replication of SARS-CoV-2. We confirmed that CMs have the predicted 120kD band when probed for with an antibody against ACE2 (Figure. 2A). By immunohistochemistry we confirmed the expression of ACE2 in the CMs, which was predominately cytoplasmic, but in some cells was detected on the cell surface. Membrane expression is highlighted by arrows in Fig. 2B left most panel. We confirmed that the cells are indeed CMs by immunostaining for Troponin T, a known marker for myofibrils (27) (Figure. 2B middle). Overlay of the ACE2 and troponin staining is shown in the left most panel of Figure 2. In MSDW 2235 patients out of the 4228 patients with troponin I values also had IL6 values. Within this group 84% of COVID-19 patients (1872/2235) have elevated IL6 values of 30 pg/ml (18) as would be expected for patients with an active infection. Numerous ILs have been shown to be elevated in COVID-19 patients (18, 28). However, since IL6 values in blood are most commonly available in clinical data sets we focused on IL6 and IL1-β. We hypothesized that studying SARS-CoV-2 infection of CMs in the presence of ILs would better approximate what myocardial tissue encounters *in vivo*. Hence, we tested the effects of SARS-CoV-2 infection on CMs in the absence and presence of ILs in the culture medium. Compared to the controls (MOCK infected cells in the absence of ILs) the CMs treated with IL-6 and IL-1β appeared larger in diameter and the myofibrils immunostained with Troponin T were disorganized (Figure 2C compare top and 2^nd^ level panels). Infection with SARS-CoV-2 at a multiplicity of infection (MOI) of 0.1 alone also appeared to increase the cell diameter compared to Mock controls (Level 3 panel). Infection in the presence of ILs produced the maximal effects as assessed by apparent increase in cell size and disorganized myofibrils (Figure 2C lowermost panel). SARS-CoV-2 at a MOI of 0.1 for 48hrs can infect CMs as confirmed by immunostaining for viral nucleocapsid protein (NP) (Figure. 2C red staining – leftmost photos, bottom two panels). All the infected cells were positive for ACE2 (green stain), and Troponin T (cyan stain) and the myofibrils were disorganized as indicated by the Troponin T staining (Figure 2C lower panels). These findings were reproduced in a second CM line as shown in Supplementary Figure 4. To determine if the observed increases in cell size were statistically meaningful we randomly measured cellular diameter in CMs without and with infection and treatment with ILs. We measured the randomly the diameter of over 50 cells from two different CM lines using Image J and found that interleukins alone significantly increased the cell diameter. Infection alone did not significantly increased the cell diameter compared to Mock controls, but there was in increase in cell diameter in the presence of ILs (Figure 2D). We also tested whether interleukins increased the percentage of cells infected by SARS-CoV-2. We stained the cells for viral NP and Hoechst nuclear stain to count total cell numbers and cells that stained for NP protein. We used the In Cell Analyzer to automate the cell count for percentage of cells positive for NP. At both 48 and 72 hours post infection, ILs did not alter the level of infection of CMs by SARS-CoV-2 (Figure 2E). From these cells we collected the media to determine the production and shedding of virus. We collected supernatants from infected CMs using three different lines, MSN-07-07S, MSN-08-06S and MSN-31-01S, and performed TCID50 analysis (an assay to quantitate viral infections) at 48 and 72hrs post-infection (Fig. 2F). We observed that all three CM lines were productively infected using a MOI of 0.1. We confirmed that ILs alone did not impact the replication of the SARS-CoV-2, as there were no significant differences in the release of viral particles with and without interleukins.

An important indicator of CM damage is the release of troponin from the cells. Hence, we measured the levels troponin I in the culture medium of cells without and with infection, in the absence and presence of interleukins. For this we used a commercial ELISA -kit and measured culture medium troponin levels from three different CM cell lines. IL treatment did not did not cause the release of troponin. When CMs were infected with SARS-CoV-2 at a MOI of 0.1 there was increased release of troponin into the culture medium but the effects were not statistically significant. However, under these conditions, infection in the presence of ILs caused a statistically significant near threefold increase in levels of troponin I in culture medium (Figure 2G). These results indicate that infection in the presence of interleukins can substantially damage cardiomyocytes, although interleukins alone or infection in the absence of interleukins can also damage cardiomyocytes. In vivo, this distinction may not be relevant.

### Effects of SARS-CoV-2 infection on beating of ventricular cardiomyocytes and left ventricular ejection fraction show similar characteristics

A well-known hallmark for healthy CMs derived from iPSC lines is their ability to beat in a coordinated manner in vitro (29). We recorded the beating of 3 different CM lines under the following conditions: MOCK infection, infection with SARS-CoV-2 with MOI 0.1, treatment with ILs alone and infection in the presence of ILs 48hrs post infection (see Movie1 related to Figure 3A). To quantitate the number of beats per minute, we counted how many beats were occurring over one minute within 5×5 grids (25 grids in total) superimposed over the video, blind to the treatment. In Figure 3A, we show a representative snapshot of the field of recording with the number of beats per minute. Control cells show coordinated beating over the entire field (Figure 3A upper left panel). Infection with SARS-CoV-2 at a MOI of 0.1 causes considerable alteration in coordinated beating such that the beating is decreased in some regions while it is near normal in several regions (Figure 3A, right panel). Treatment with interleukins alone causes similar effects although in all regions we see a decrease in beat frequency (Figure 3A lower left panel). Infection in the presence of interleukins results in a marked decrease in beating with no beats in nine of the 25 fields. (Figure 3A bottom right panels) Summary statistics across the multiple cell lines show that both interleukins and infection decrease beating (4.17 and 4.22 respectively vs. 9.3 beats/min for control) with a more pronounced decrease (2.08 beats per min) when infection occurs in the presence of interleukins, which is likely to be the most physiologically relevant scenario (Figure 3B).

Since the beating of CMs in culture is broadly indicative of myocyte contraction *in vivo* we reasoned that alterations in contractility during COVID-19 in patients could result in a decrease in left ventricular ejection fraction (LVEF), a physiological measure routinely used in clinical diagnosis. Hence, we looked for COVID-19 patients without prior cardiac disease who had elevated troponin levels and showed decreased LVEF. Using data from two Mount Sinai health System hospitals we identified a cohort of 206 COVID-19 patients without prior heart disease for whom imaging data along with troponin I and IL6 levels were available. In Figure 3 C we show a plot of COVID-19 patients with normal (Black dots) and reduced LVEF (blue squares) in terms of their relationship to blood IL6 and troponin I levels. We find that nearly 50% (20/41) of the patients with LVEF<51 had troponin levels above 1, while for normal LVEF it is 22/165 (13.3%). We also find that 6/41 (14.6%) patients have troponin I levels greater than 20 ng/ml (ln of 3), and none of the normal LVEF patients are above 20. No relationships between IL levels and LVEF status could be readily observed. Representative cardiac images from COVID-19 patients are shown in Figures 3C-F. These are apical views after administration of an ultrasonic enhancing agent for transthoracic echocardiogram (TTE). A COVID-19 patient with no known cardiac disease and normal LV function is shown in Movie 2, where no detected abnormalities are found in the diastolic and systolic frames. Clips of the diastolic and systolic frames are shown in Fig. 3D. The end-diastolic (EDV) and end-systolic volumes (ESV) are 104ml and 35ml, respectively with a left ventricular ejection fraction of 66%. Two patients with significantly elevated levels of blood troponin and reduced LV function (LVEF<51%) are shown in clips in Figures 3E and 3F and in Movies 3 and 4. One patient had a mid infero-lateral wall hypokinesis, but preserved LV ejection fraction (see Fig. 3E and Movie 3). The regional wall motion abnormality (RWMA) is pointed out in the red oval in Fig. 3E. The EDV and ESV are 102ml and 50ml, respectively with LVEF of 51%. The second patient with newly reduced LV function had global LV dysfunction. From the clips in Fig. 3F and Movie 4, the findings are consistent with diffuse LV wall hypokinesis and mildly decreased LVEF, EDV and ESV are 160 and 90mls respectively with LVEF of 44%. Overall the relationship between elevated troponin I level and decreased LVEF is highly significant in COVID-19 patients (Figure 3G).

## Discussion

As a respiratory virus, the multiorgan pathophysiology of SARS-CoV-2 could not entirely be predicted. Although it has been long known that ACE2, the receptor for members of the SARS family, is widely distributed, it is not clear why SARS-CoV-2 shows a range pathophysiologies including loss of smell, heart disease, kidney disease, and neurological disorders in addition to the expected severe respiratory disease. It is also not clear if these pathophysiologies of multiple organs occur in the same patients or if different, groups of patients have a propensity for different types of organ pathophysiologies. We have decided to study cardiac dysfunction in COVID-19 in relative depth as we had appropriate model systems such as healthy human subject iPSC-derived cardiomyocytes available for *in vitro* studies. Analyses of the clinical data from the hospitals of the Mount Sinai Health System indicates that significant numbers (> 10%) of COVID-19 patients develop cardiac dysfunction during infection If these ratios hold worldwide, then nearly 5 million individuals would have the potential for cardiac dysfunction that could lead to long term heart disease. The data from long-term studies of 9/11 responders indicate that this is likely to be the case (30, 31). Hence, understanding the origins of cardiac dysfunction in COVID-19 patients will be important for clinical management and therapeutic strategies to mitigate the development of heart disease when treating patients at Centers for Post-COVID-Care such the one in the Mount Sinai Health System.

Myocardial damage can occur due to multiple reasons including decrease in oxygen levels due to ARDS, the formation of thrombi as has been observed in COVID-19 patients (15), and direct injury of cardiomyocytes by viral infection and the inflammatory state of the COVID-19 patients. Our *in vitro* cardiomyocyte studies indicate that that direct injury due to infection is likely to be one of the causes of myocardial injury in COVID-19 patients. Two aspects of our *in vitro* studies are noteworthy: first, measurable cardiomyocyte injury, such as release of troponin into the culture medium, and near complete disruption of coordination and inhibition of beating, is greatest when viral infection occurs in the presence of interleukins, a scenario that is most likely similar to that seen in vivo; and second interleukins do not increase the number of cells infected which remains stable around 35%. Taken together these two observations suggest that cell communication between cardiomyocytes and other cell biological mechanisms may underlie the effects of SARS-CoV-2 infection in myocardial tissue. Such potential mechanisms merit detailed studies under conditions that consider genomic determinants that could control the pathophysiological responses. Studies await the development of animal models of COVID -19 relevant to human disease.

In conclusion by integrating analyses of clinical data and observations from cell level experiments in culture, using qualitative matches between biomarkers like troponin and physiological functions like beating we show that direct infection of cardiomyocytes can be one mechanism of myocardial injury during COVID-19.

## Supporting information

Full Supplement

Movie 1 - beating cardiomyocytes

Normal Echocardiogram

RWMA Echocardiogram

Diffuse Echocardiogram

## Data Availability

All data is available in the main text or in the supplementary materials.

## Acknowledgements

We like to thank Som Prabha for her assistance with maintaining CM lines.

## IRB-Approval

IRB approval at Icahn School of Medicine at Mount Sinai, Study-20-01561 – Myocardial injury in COVID-19.

## Author contributions

MMS, ATC, LM, ASY, AG-S and RI developed the conceptual ideas and experiments for this paper

MMS contributed to all experiments and analysis and took the lead in coordinating the study

ATC prepared all human IPSC-derived cardiomyocytes with assistance of JH, conducted the Troponin-I ELISA, and contributed to immunostaining, INcell analysis and cardiomyocyte beating.

ASY analyzed all MSDW patient data and executed machine learning analysis

LM, TK, KMW and AC performed all infections with SARS-CoV-2 under BSL3 containment

LM and TK performed TCID50 and collected the supernatants for Troponin detection under BSL3 containment

LM and KB recorded live cell beating of the cardiomyocytes under BSL3 containment

SN and PK extracted the data and curated the Mount Sinai Data Warehouse (MSDW) anonymized clinical data set.

PM organized the movies and helped with video analysis of the beating of cardiomyocytes

ND and CS developed and characterized the iPSC lines used in the studies

RET and BH assisted with cardiomyocyte experiments and did blind counting of beating of cardiomyocytes

AS and IT conducted INcell analysis for quantification of infected cardiomyocytes

RI-K, KC NK, GG, EA, MM, SL, JN obtained, organized and analyzed the cardiac imaging data.

MMS, ASY, JG and RI wrote the paper and all authors reviewed the manuscript and provided comments.

“All data is available in the main text or in the supplementary materials.”

## List of Supplementary Materials

**Figure S1. Data driven assessment to determine optimal manner dealing with missing data during the model development process**.

**Figure S2. Results of recursive feature elimination during the model development phase using preoperative only dataset**.

**Figure S3. The statistical comparison of two classification algorithms performances in the form of Critical Difference (CD) plots**.

**Figure S4** – **CM line MSN08-06S can be infected by SARS-CoV-2 and have disrupted Troponin T.**

**Figure S5: Interleukins do not increase the infectivity of SARS-CoV-2 in 4 different CM lines tested**.

**Table S1. Descriptive characteristics of the overall development (80%) dataset, and 20% test dataset**.

**Table S2: Missing value number of development set and test set**.

References 32-41 are in the Supplementary only.

## List of Movies

### Please go to https://iyengarlab.org/ to see the Movies

Movie 1 – Live cell beating of hIPSC cardiomyocytes at 48hrs post infection with SARS-CoV-2. The movie has been sped up 6X and displays MSN08-06S CM line beating in the BSL3 in the following order: CM line MSN08-06S (8-48h): MOCK, Infection only at MOI 0.1, Interleukins (ILs) only (IL-6 and IL-1β each at 30ng/ml) and ILs with Infection.

All patients are confirmed to be COVID-19 positive and no history of cardiac disease. Movie 2 has normal LVEF, while 3 and 4 have newly reduced LVEF<51.

Movie 2 - Normal demonstrates an apical 4-chamber view (after administration of an ultrasonic enhancing agent) from a transthoracic echocardiogram. The findings are consistent with preserved left ventricular ejection fraction and no regional wall motion abnormalities. The end-diastolic and end-systolic volumes are 104ml and 35ml, respectively with a left ventricular ejection fraction of 66%.

Movie 3 – Regional Wall Motion Abnormality (RWMA) demonstrates an apical 3-chamber view (after administration of an ultrasonic enhancing agent) from a transthoracic echocardiogram. The findings are consistent with basal and mid infero-lateral wall hypokinesis despite preserved left ventricular ejection fraction. The end-diastolic and end-systolic volumes are 102ml and 50ml, respectively with a left ventricular ejection fraction of 51%

Movie 4 – Diffuse demonstrates an apical 4-chamber view (after administration of an ultrasonic enhancing agent) from a transthoracic echocardiogram. The findings are consistent with diffuse left ventricular wall hypokinesis and mildly decreased left ventricular ejection fraction. The end-diastolic and end-systolic volumes are 160ml and 90ml, respectively with a left ventricular ejection fraction of 44%.

