## Supplementary material for "Physiology of cardiomyocyte injury in COVID-19": Full Supplement

#### Abstract

COVID-19 affects multiple organs. Clinical data from the Mount Sinai Health System shows that substantial numbers of COVID-19 patients without prior heart disease develop cardiac dysfunction. How COVID-19 patients develop cardiac disease is not known. We integrate cell biological and physiological analyses of human cardiomyocytes differentiated from human induced pluripotent stem cells (hiPSCs) infected with SARS-CoV-2 in the presence of interleukins, with clinical findings, to investigate plausible mechanisms of cardiac disease in COVID-19 patients. We infected hiPSC-derived cardiomyocytes, from healthy human subjects, with SARS-CoV-2 in the absence and presence of interleukins. We find that interleukin treatment and infection results in disorganization of myofibrils, extracellular release of troponin-I, and reduced and erratic beating. Although interleukins do not increase the extent, they increase the severity of viral infection of cardiomyocytes resulting in cessation of beating. Clinical data from hospitalized patients from the Mount Sinai Health system show that a significant portion of COVID-19 patients without prior history of heart disease, have elevated troponin and interleukin levels. A substantial subset of these patients showed reduced left ventricular function by echocardiography. Our laboratory observations, combined with the clinical data, indicate that direct effects on cardiomyocytes by interleukins and SARS-CoV-2 infection can underlie the heart disease in COVID-19 patients.

#### Materials and Methods:

##### Study Approval

This study was approved by the Institutional Review Board (IRB) of the Icahn School of Medicine at Mount Sinai and complies with all institutional and federal requirements regarding research using data from human subjects.

##### Clinical Data & study population

This retrospective analysis was conducted using de-identified electronic medical record (EMR) data from patients diagnosed with COVID-19 positive within the Mount Sinai Hospital System, between 9 March and July 15, 2020. The Mount Sinai Health System is a network of 8 hospital campuses and over 400 ambulatory practices spanning the New York metropolitan area (17). COVID-19 diagnosis was based on positive polymerase chain reaction (PCR)-based clinical laboratory testing for the SARS-CoV-2 virus. Patient's data at Mount Sinai Health System were collected by Mount Sinai Data Warehouse team and de-identified data were released for research purposes.

All collected data and events occurring during the time that medical attention was provided to the patient during the visit were defined as an encounter data. For patients with more than one documented encounter in the data set, such as those with a readmission or transfer from another affiliated facility, only the first encounter data was included for analysis. Patient characteristics were collected from the EMR. Basic demographics included age, sex, race, ethnicity, body mass index (BMI), smoking status (current smoker, former smoker, never smoker, passive smoker). Patients vitals at admission (Temperature maximum, systolic blood pressure min, Oxygen saturation minimum, heart rate etc.) its maximum/minimum values recorded during the stay and comorbidities present on admission (e.g., cancer, hypertension, heart failure, cardiac arrhythmias, cerebral infraction, chronic kidney disease, acute kidney injury, HIV/AIDS, liver disease, diabetes, etc.) were also collected from the EMR. Lab data of patients collected several times during the encounter, maximum recording collected for all lab features except few with minimum values (Albumin, EGFR, Hemoglobin, Neutrophil (%), Hematocrit, Platelet, Ferritin) based on its severities'.

Under an agreement with the IRB, which is a precursor and separate from our study, the Mount Sinai Data Warehouse released de-identified clinical data of all patients with COVID-19 who had or were having treatment in the Mount Sinai Health System. Since de-identified data were used, patient consent was not sought. We did univariate significance analyses of the differences in continuous features between patients without cardiac disease and patients with cardiac disease using Student's t test and of categorical features using the $\chi^2$ test in all the resultant datasets.

Raw Data from March 9$^{th}$ to July 15$^{th}$ acquired from Mount Sinai Data Warehouse (MSDW) and followed preprocessing steps as described below.

**Cardiac Imaging Data**

For Cardiac point of care ultrasounds/echocardiograms they were performed by certified emergency room physicians and critical care physicians on patients when deemed necessary by the attending physician. Formal echocardiograms were done on those for whom further information necessary. The patients were identified retrospectively, and subsequent data collection occurred to obtain baseline characteristics, prior medical history including cardiovascular disease history, hospital outcomes of mortality and intubation, anthropometric measurements, and laboratory values.

**Analyses of MSDW Data**

**Data Pre-processing**

We applied the following basic pre-processing steps to the raw clinical data in the development set:

1. Only records for patients with a positive SARS-CoV-2 test were retained, and the rest eliminated.
2. Only the first record of encounter data among all those with the same patient ID is retained, and the rest eliminated.

3. Vital sign features minimum and maximum values (e.g. systolic blood pressure min & systolic blood pressure max) retained during the course of encounter.
4. Lab data features maximum values retained during the encounter except few features minimum values were calculated of (Albumin, EGFR, Hemoglobin, Neutrophil (%), Hematocrit, Platelet, Ferritin)
5. Merged (encounter data, vital signs data & lab data) on patient ID.
6. Features with missing values in more than 60% of the patient records (seven in this case) were dropped to prevent their adverse effects on data analyses.
7. Any of the categorical features, e.g., sex and race, that had exactly the same value for over 99% of the patients, i.e., were effectively constant, were removed, since they were unlikely to be predictive. The values of the remaining categorical features were converted into numerical form (1, 2, 3…) based on the order of the discrete values appearing in the data. Undefined or unclear entries for these features were recorded as missing values.
8. The continuous features, e.g. AGE, O2SAT_min, were normalized by converting them into respective z-scores by applying the formula $\frac{X-\mu}{\sigma}$, where $X$, $\mu$ and $\sigma$ are the value, mean and standard deviation of a given feature.

The same pre-processing steps were applied to test set. In the steps where the pre-processed values of the features were dependent on those determined from the development set, e.g., numerical values of categorical features in step 7, and mean and standard deviation in step 8, the same values determined from the development set were applied to the test set. This was necessary to ensure that a predictor trained on the development set is applicable to the test set. It also had the desirable effect of the steps not being dependent on the relatively smaller sizes of the test set, which may make difficult to estimate parameters like mean and standard deviation in step 8 reliably.

The resulting development and test set were used for the machine learning analyses described in the following subsections.

**Development and evaluation of predictive machine learning models**

For predictor construction, we further randomly split the development set into training and holdout subsets of sizes 75% (n=2536) and 25% (n=846), respectively. The analysis steps used in this process, specifically missing value imputation and feature selection (described below), were used to learn candidate predictors from the training set, which were in turn evaluated on the corresponding holdout set. The logistic regression (LR) as implemented with its default parameters in Python's scikit-learn library (32), as well as the and eXtreme Gradient Boosting (XGBoost) algorithm (33), as implemented with its default parameters in Python's XGBoost package, were used to develop these candidate predictors. This process was repeated a hundred times. The performance of the candidate predictors was quantified as the average value of their evaluation measure in each round. The overall predictor development process has followed as shown in Figure 1 **C-I**.

All predictor development and validation were conducted using area under the receiver operating characteristic (ROC) curve (AUC score) (34) AUPRC – area under the precision recall curve; P_max – precision maximum; R_max – recall maximum; F_max – f1-score which is harmonic mean of precision and recall; Accuracy as the evaluation metric.

##### Missing value imputation

Several features in our dataset had missing values (**Table S2**). To maximize feature inclusion in the predictive modeling process, we evaluated the impact of imputing and including missing values among features (35). Specifically, we considered an increasing number of features with increasing levels of missing values (≤ 0%, 5%, 10%, …, 60%) in the training subset of the development set. Then, we replaced the missing values for each feature in the development set with its mean or mode in the training subset, depending on whether the feature was continuous or discrete, respectively. Finally, we trained and evaluated candidate predictors using the two algorithms listed above on the training and holdout subsets of the development set respectively. Using the results, we estimated the level of missing values that would be the most beneficial to be included and imputed.

##### Feature selection

In predictive modeling with clinical data, several features may be irrelevant to the target problem, and several may be redundant with other features, making accurate prediction challenging (36). To address this challenge, and to develop a predictor more parsimonious than one based on all features, we utilized established feature selection methods (36). Specifically, we adopted the same training-holdout setup and classification algorithms as described in the previous subsection, and assessed how well features selected automatically using the Recursive Feature Elimination (RFE) algorithm (37) performed. This algorithm ranks the full set of features and then evaluates the performance of a classifier trained using one of our two candidate algorithms on the *k* top-ranked features, where values of *k* are pre-specified. Based on the evaluation results, RFE infers the value of *k*, and as many features, that are likely to perform the best with the given algorithm. The candidate values of *k* we tested within RFE were 1, 5, 10, 15, 20, 25, 30, 40, 50, 60, 70 and 86. For the both classification algorithm and RFE-inferred value of *k*, we selected the final set of features as the top *k* highest-frequency features selected by RFE for that algorithm across the 100 runs of the training-holdout setup.

**Final predictor development and validation**

Once we had determined the acceptable level (% of patients) of missing values, and the features satisfying this requirement, we imputed the corresponding missing values in test set using mean or median values in the development set. Next, we trained and evaluated the final COVID-19 mortality predictor on the imputed development and validation sets respectively. The predictor was trained using the both classification algorithms (XGBoost & Logistic Regression) on the top 15 selected features and the associated classification algorithm determined as described in the previous subsection. The purpose of evaluating both of these predictors was to assess how their performances generalized to an independent patient population.

**Biosafety**

All experiments involving SARS-CoV-2 were conducted under BSL3 containment in accordance to the biosafety protocols developed by the Icahn School of Medicine at Mount Sinai.

**Cells and viruses**

The SARS-CoV-2 USA-WA1/2020 strain, isolated from an oropharyngeal swab from a patient with a respiratory illness who developed clinical disease (COVID-19) in January 2020 in Washington, USA, was obtained from BEI Resources (NR-52281). The virus was propagated in Vero E6 cells. Viral stocks were grown in Vero E6 cells as previously described, and were validated by genome sequencing (38).

Vero E6 (ATCC, CRL-1586) were maintained in DMEM (Corning) supplemented with 10% FBS (Peak Serum) and Penicillin/Streptomycin (Corning) at 37°C and 5% CO2. All cell lines used in this study were regularly screened for mycoplasma contamination using the Universal Mycoplasma Detection Kit (ATCC, 30-1012K).

**Healthy human subject iPSC-derived cardiomyocytes**

The iPSC lines used in this study was obtained a library of 40 lines from healthy human subjects (25) Four iPSCs (MSN07-07S, MSN08-06S, MSN31-01S, and MSN39-04S) were maintained in StemFlex media (Gibco) on hES-qualified Matrigel (Corning)-coated 6-well cell culture plates at 37°C with 5% CO2. hiPSCs were passaged every 3 days and supplemented with Rock Inhibitor Y-27632 (SelleckChem) for 24 hours after each passaging. For cardiomyocytes differentiation (39), hiPSCs were treated with 10 µM GSK3 inhibitor CHIR99021 (SelleckChem) in RPMI1640 (Gibco) supplemented with B27 without insulin (Gibco) for 24 hours. After an additional 48 hours, media was changed and then supplemented with 5 µM IWR1 (Sigma) for 48 hours. From day7 onwards cells were cultured in RPMI1640 medium supplemented with B27 with insulin (Gibco) and media was changed every 3 days. Beating was generally observed around day 8-10. Between days 15-23, iPS-cardiomyocytes were washed with DPBS (Gibco) and cultured in RPMI1640 without glucose supplemented with 4 mM sodium lactate (Sigma) and 25 mM HEPES (Gibco) for metabolic switch purification (40). Lactate media was refreshed every 4 days. After day 30, iPS-cardiomyocytes were replated into assay plates for use in experiments. Typically, the cells were maintained for another 15-30 days before being subjected to treatment. Culture media was changed every three days,

Infection of human cardiomyocytes were carried out at a MOI1 of 0.1 unless indicated otherwise. All experiments involving live virus was carried out under BSL3 containment. After the experiments the virus was inactivated and cells fixed before the plates were removed from the BSL3 for staining and further analyses.

All live cell beating recordings were made on an EVOS M5000 (Thermo) in the BSL3 containment facility. The cell lines used for these physiological experiments had been in culture for 45-60 days. Movies were made for one-minute recordings for 3-4 different fields per condition. These movies had a 5x5 grid (25 boxes) juxtaposed over the video image. We blindly counted the number of beating cells per box for all conditions.

All microscopy was performed on a Zeiss LSM 880 confocal microscope. Cell diameter measurements were performed using the LSM acquired images that were imported into ImageJ.

**Troponin I ELISA**

Human Troponin I ELISA Assays were was performed using a commercially available kit (Thermo Fisher Catalog # EHTNNI3). Typically 100µls of Cell culture medium was used for the assay. We followed the manufacturer's protocols without any modification.

**Measurement of virus levels- TCID50 (50% Tissue Culture Infectious Dose) assay**

TCID50 assays were performed as described previously (41) In brief, Vero E6 cells were infected with serial ten-fold dilutions of the supernatants harvested from cells treated with IL-6 and/or IL-1β. On day 4 post-infection, cytopathic effect (CPE) was assessed by crystal violet staining, and TCID50 was calculated using the Reed & Muench method (41).

**Reproducibility and Statistics:**

All experiments were carried out in at least 3 replicates and the findings were reproduced in 3 to 4 separate experiments. All averages are from at least 3 separate experiments. Except for the CM cell beating experiments, we used three different cell lines in triplicated and imaged several different areas of each well for beating. This was done for two separate experiments for all 3 CM cell lines. Averages are from the two different experiments.

We used two-tailed ANOVA with Bonferroni corrections for multiple comparisons or t-test analysis.

**Cardiac Imaging:**

Echocardiographic imaging was done as part of the clinical care of COVID-19 patients as determined by their physicians. The studies were performed using portable ultrasound machines: CX50, Philips Medical Systems, Bothell, Washington and Vivid S70, General Electric Medical Systems, Milwaukee, Wisconsin. Contrast-enhanced echocardiograms were performed using contrast-specific, low mechanical index settings after administration of ultrasonic enhancing agents, either Definity (perflutren lipid microsphere), Lantheus Medical Imaging or Optison (perflutren protein type-A microspheres), GE Healthcare. Echocardiograms for COVID19 positive patients were performed in compliance with the following specific measures. Some studies were performed with the Transthoracic echocardiography (TTE) using the EPIQ echocardiography system (Philips North America, MA, USA). TTE studies were performed using a focused examination protocol according to American Society of Echocardiography recommendations for use of echocardiography in patients with confirmed COVID-19 infection.

**Software**

The following Python version and packages are used in model building.

Python 3.6.7

Pandas 0.24.2

NumPy 1.16.4

Scikit-learn 0.21.2

Scipy 1.3.1

Matplotlib 3.1.0

seaborn 0.9.0

xgboost 0.80

MATLAB 9.5.0.944444 (R2018b)

### Supplementary Figures

**Figure S1**

**A**

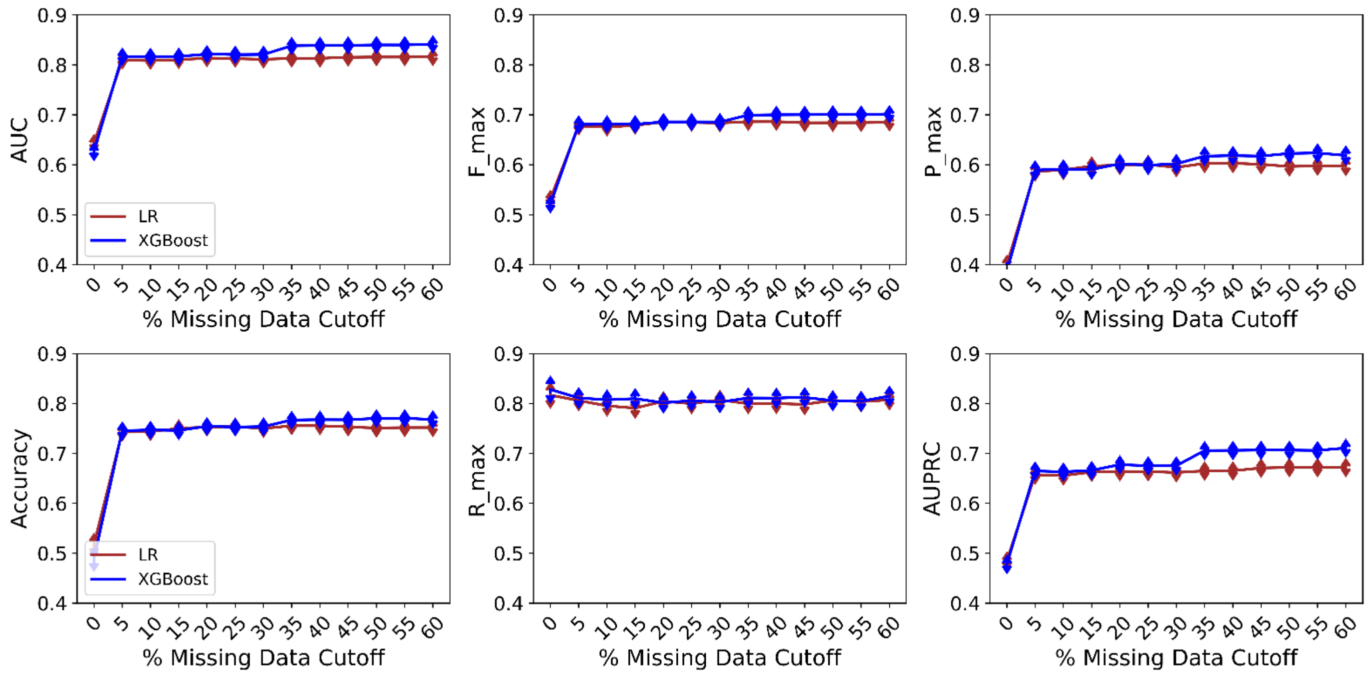

**B**

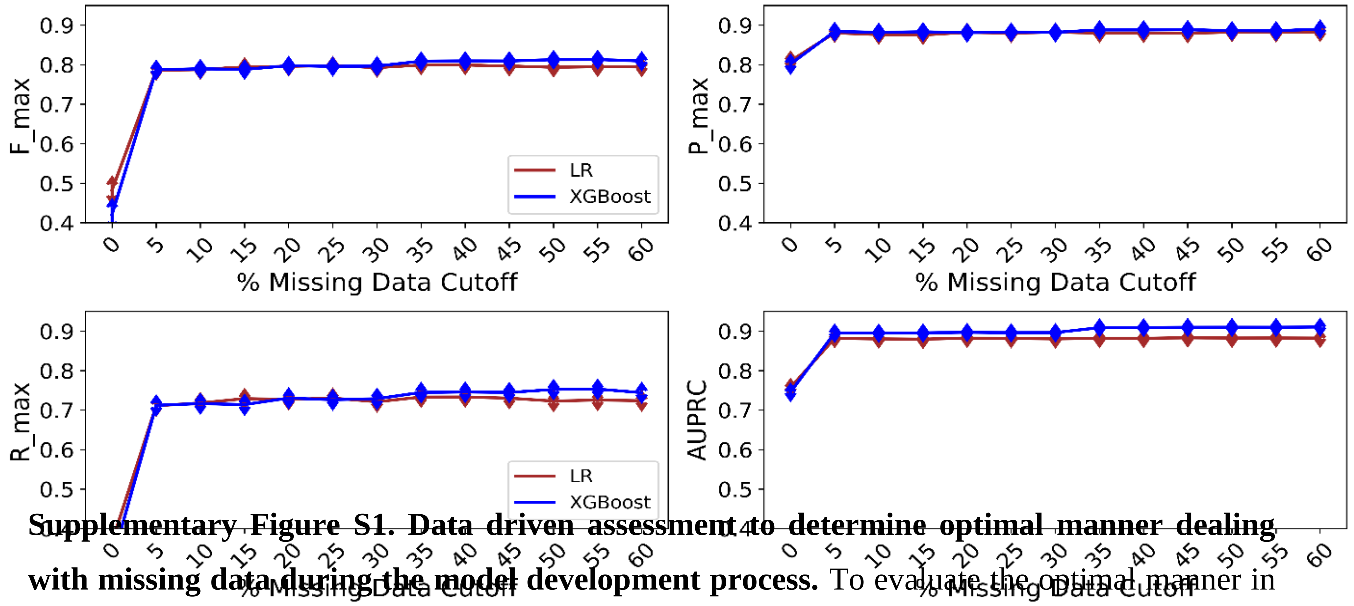

**Supplementary Figure S1. Data driven assessment to determine optimal manner dealing with missing data during the model development process.** To evaluate the optimal manner in which to deal with features with missing data, different cutoffs were taken in 5% increments (0%

to 60%) of missing data and using the training set, the mean-mode imputation method was evaluated (mean for continuous features, mode for categorical features). Two different classifiers (LR, XGBoost) were then trained at each of the percent missing data cutoffs on the training dataset and validated on the 20% validation dataset with subsequent evaluation by six different metrics (AUC, Accuracy, F_max, R_max, P_max, AUPRC) in order to assess model performance at each cutoff. Each model was run 100 times for each missing data cutoff and the average of the performance was plotted for each metric for the **A.)** Troponin level more than 0.09 and **B.)** Troponin level less than equal to 0.09 (AUC and Accuracy are not presented as these are class independent metrics). Data points show the average AUC score for each candidate algorithm and missing value level, with error bars shown by whiskers.  AUC – area under the receiver operating characteristic curve; AUPRC – area under the precision recall curve; LR – logistic regression; P_max – precision maximum; R_max – recall maximum; F_max – f1-score which is harmonic mean of precision and recall ;XGBoost – extreme gradient boosting.

**Figure S2**

**A**

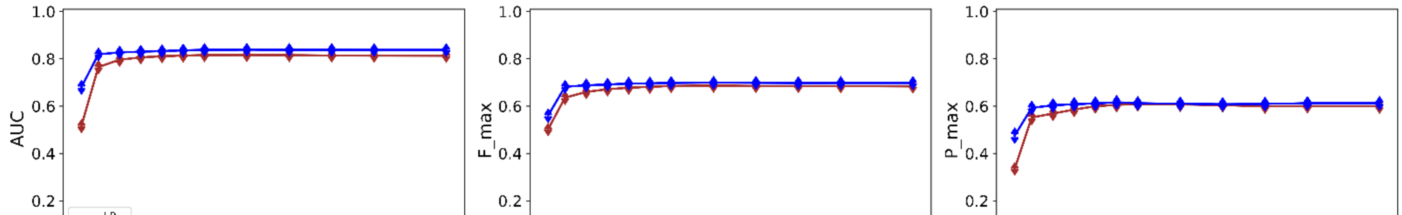

**B**

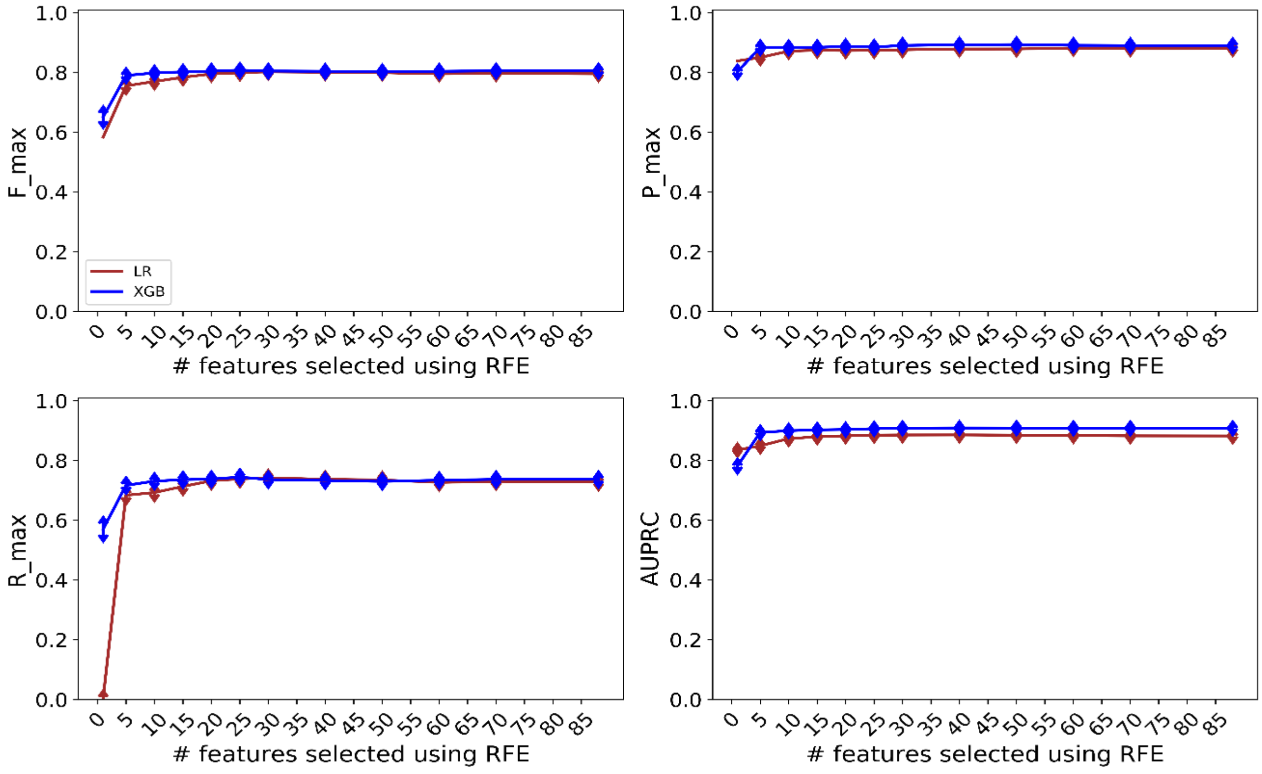

**Supplementary Figure S2. Results of recursive feature elimination during the model development phase using preoperative only dataset.** Recursive feature elimination is a feature selection method that fits a model and removes the weakest feature (or features) until the pre-specified number of features is reached. Various incremental percentages (0% to 100%) of the overall number of features (100% features = 86 features) were assessed. Two different classifiers (LR, XGBoost) were then trained at each of the percent missing data cutoffs on the training dataset 75% and validated on the 25% validation dataset with subsequent evaluation by six different metrics (AUC, Accuracy, Fmax, Rmax, Pmax, AUPRC) in order to assess model performance at each cutoff. Each model was run 100 times for each missing data cutoff and the average of the performance was plotted for each metric for the **A.)** Troponin level more than 0.09 and **B.)** Troponin level less than equal to 0.09 (AUC and Accuracy are not presented as these are class independent metrics). Data points show the average AUC score for each candidate algorithm and missing value level, with error bars shown by whiskers. AUC – area under the receiver operating characteristic curve; AUPRC – area under the precision recall curve; LR – logistic regression P_max – precision maximum; R_max – recall maximum; F_max – f1-score which is harmonic mean of precision and recall; XGBoost – extreme gradient boosting.

**Figure S3**

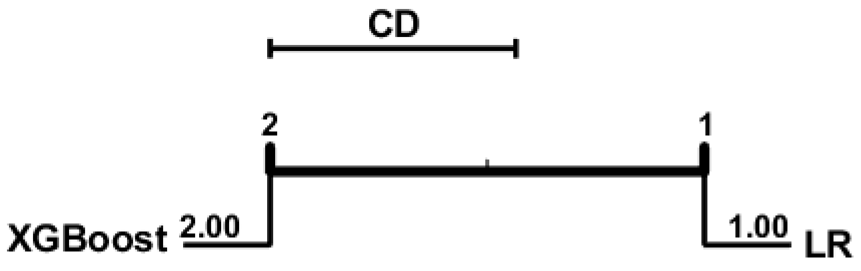

**Supplementary Figure S3. The statistical comparison of two classification algorithms performances in the form of Critical Difference (CD) plots:** Classification algorithms, represented by (vertical + horizontal) lines, are displayed from left to right in terms of the average rank obtained by their resultant models in each of the 100 simulations on validation data set. The classifiers producing statistically significantly different performance because both classifiers are not connected by horizontal lines. These results show that the XGBoost is the best performer overall. The CD plots were drawn using open-source Matlab code.

Supplementary Figure 4

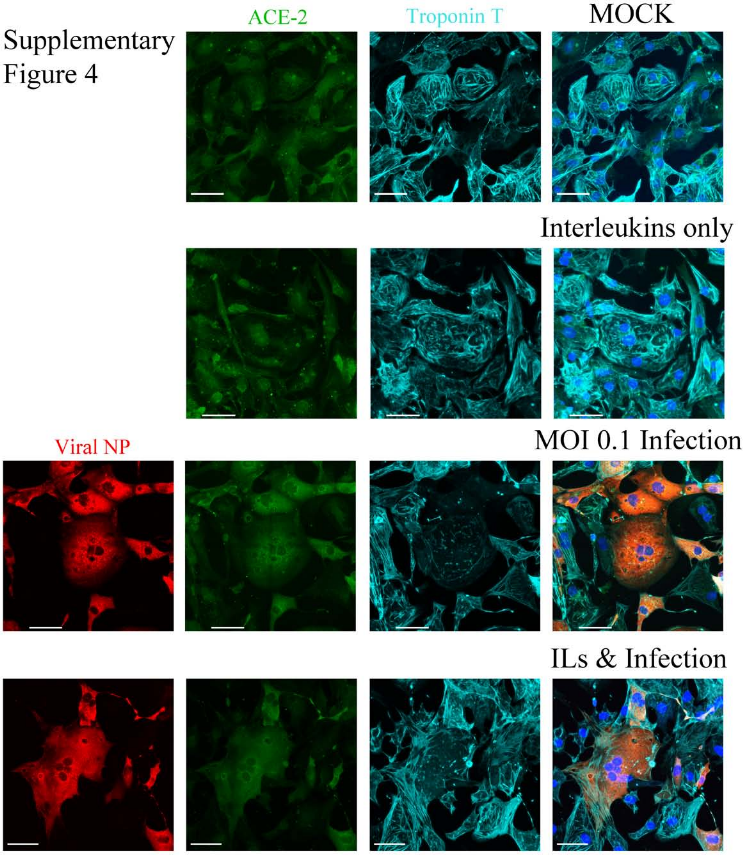

**Figure S4** - Mock infected cardiomyocytes express ACE-2 and Troponin T, nuclei are stained by Hoescht. CM treated with 30ng/mls of IL-6 and IL-1β still express ACE-2 and the cells appeared larger, but the Troponin T appeared more disorganized. (C) CM can be infected with SARS-CoV-2 with a MOI 0.1 as detected by positive immunostaining for viral NP (Red), all infected cells were positive for ACE-2 with some cells having notable amounts of Troponin T disruption. ILs with infection with SARS-CoV-2 did infect the cells, with considerable increases in cell diameter and more Troponin disruption. The scale bar for all images is 50µm. This experiment using line MSN08-06S is a repeat of experiment shown in Figure 2C and are the cells in the CM beating in Movie 1.

### Figure S5

A.

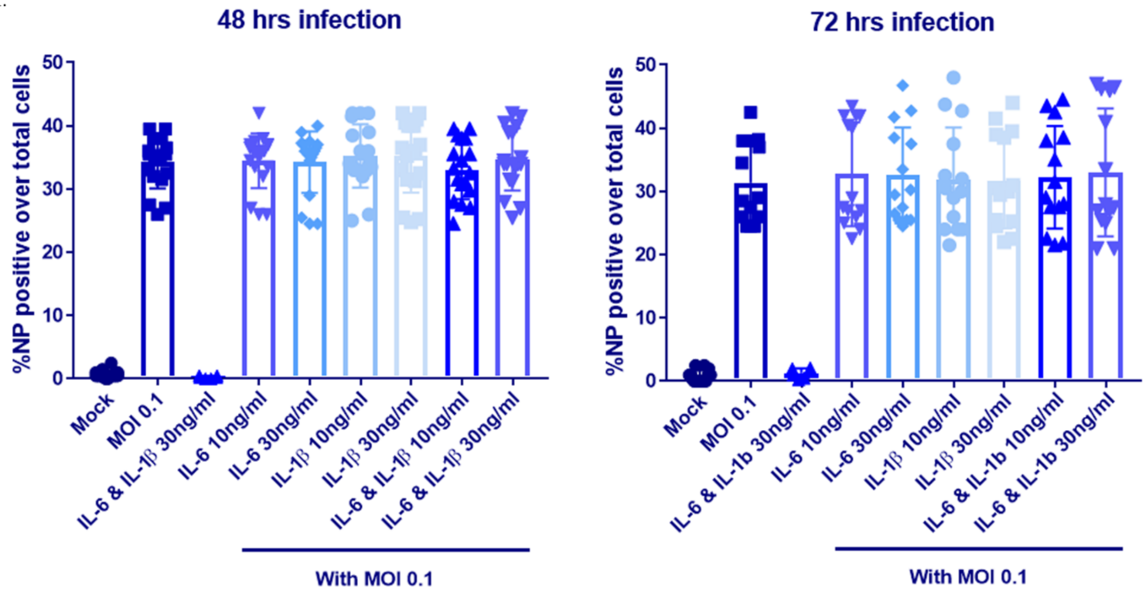

B.

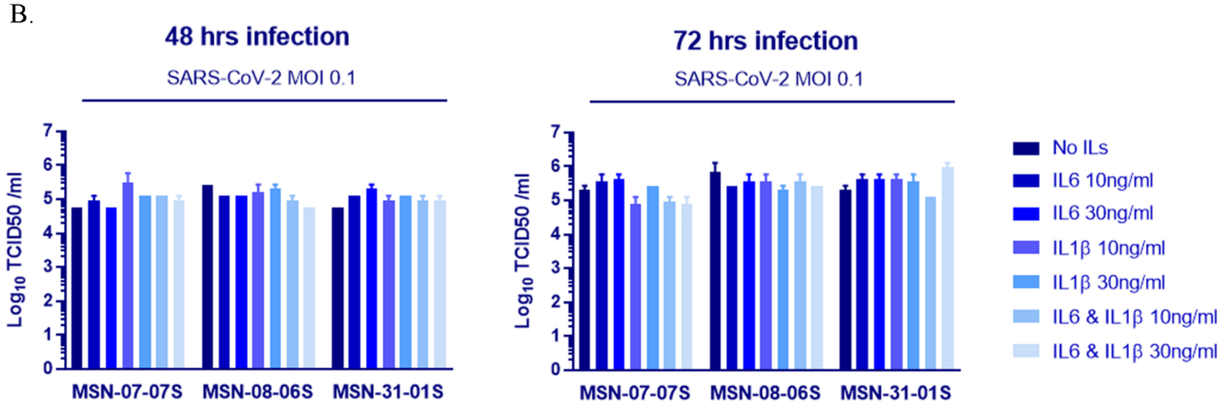

**Figure S5: Expanded version of main figures 2E and F. A.** We counted the numbers of cells stained by N protein antibody as a measure of infectivity, using either IL-6 and/or IL-1β at 10 or 30ng/ml. Each point is representative from one well of a 96 well plate with 10,000 CM cells. The bar graph is the average of four different CM lines measuring viral NP protein positive cells over total number of cells. Counting was done in an automated fashion using the InCell Analyzer. No statistical difference was determined between infection alone or in any combination of ILs tested. **B.** ILs do not increase the percentage of CM cells infected by SARS-CoV-2. To confirm that CM were being productively infected and shedding SARS-CoV-2 into the culture media, we collected supernatants from 3 different CM lines 48 and 72hrs post-infection, with or without ILs and performed a TCID50 plaque assay. We observed that CM lines are infected and the level of infection, as assessed by virus release in to culture medium did not increase with the addition of IL-6 and IL-1β, there was no statistical difference detected for any condition when comparing to No ILs.

**Table S1. Descriptive characteristics of the overall development (80%) dataset, and 20% test dataset.** Characteristic table for development set and test set are presented. Both data sets further divided patients with and without cardiac disease. For continuous variables shown with mean and standard deviation in parentheses. Counts and percentages (in parentheses) are presented for categorical variables.

**Table S2: Missing value number of development set and test set.**

Data acquired from MSDW and many features have missing values which are listed in this table. It represents the number of missing values in development set and test set that are imputed by missing value imputation techniques in the model.

| TABLE S1 | Development Set (n = 3382) , 80 % of July 15th | |
|---|---|---|
| **Variables (Continous Variables)** | **Total (n=3382)** | **Without Cardiac Disease (n = 2510)** |
| AGE | 68.39 (14.67) | 66.69 (14.97) |
| ALBUMIN | 2.47 (0.71) | 2.47 (0.72) |
| ALT | 148.88 (506.51) | 155.88 (533.02) |
| ANION_GAP | 16.9 (5.56) | 16.82 (5.66) |
| AST | 259.19 (1114.6) | 271.19 (1192.09) |
| BASOPHIL (%) | 0.78 (0.88) | 0.76 (0.86) |
| BMI | 28.48 (7.48) | 28.45 (7.53) |
| BRAIN NATRIURETIC PROTEIN | 513.78 (1381.68) | 308.59 (1012.71) |
| BUN | 55.96 (44.61) | 53.69 (44.76) |
| C-REACTIVE PROTEIN | 189.23 (121.25) | 194.3 (122.47) |
| CALCIUM | 8.92 (0.9) | 8.92 (0.9) |
| CHLORIDE | 109.97 (8.85) | 110.07 (8.84) |
| D-DIMER | 5.47 (5.44) | 5.63 (5.55) |
| DIASTOLIC_BP | 73.92 (15.17) | 74.12 (14.81) |
| DIASTOLIC_BP_MIN | 48.36 (15.83) | 49.13 (15.94) |
| DIASTOL_BP_MAX | 96.81 (17.48) | 96.14 (17.3) |
| EGFR | 35.44 (28.9) | 37.45 (30.48) |
| EOSINOPHIL # | 0.2 (0.39) | 0.2 (0.42) |
| EOSINOPHIL (%) | 2.27 (3.35) | 2.18 (3.3) |
| FERRITIN | 1139.88 (1865.7) | 1169.3 (1913.5) |
| GLUCOSE | 274.63 (204.43) | 272.5 (209.56) |
| HCO3 VENOUS | 26.53 (6.02) | 26.29 (6.08) |
| HEART_RATE | 95.66 (20.67) | 97.08 (20.09) |
| HEART_RATE_MAX | 120.21 (26.38) | 119.84 (25.67) |
| HEMOGLOBIN | 10.56 (2.81) | 10.71 (2.82) |
| INR | 1.57 (1.15) | 1.49 (0.93) |
| INTERLEUKIN-6 | 659.3 (7774.61) | 771.59 (8948.25) |
| LDH | 751.96 (1181.87) | 772.7 (1296.4) |
| LYMPHOCYTE # | 1.71 (2.37) | 1.74 (2.55) |
| LYMPHOCYTE (%) | 21.06 (12.0) | 21.14 (12.17) |
| MCH | 30.6 (2.87) | 30.61 (2.83) |
| MCHC | 33.61 (1.47) | 33.66 (1.49) |
| MCV | 93.22 (7.76) | 93.03 (7.63) |
| MEAN PLATELET VOLUME (MPV) | 9.61 (1.68) | 9.53 (1.69) |
| MONOCYTE # | 0.89 (0.65) | 0.88 (0.68) |
| MONOCYTE (%) | 10.16 (5.27) | 10.0 (5.16) |
| NEUTROPHIL # | 21.2 (94.88) | 22.31 (109.32) |
| NEUTROPHIL (%) | 66.42 (15.79) | 66.55 (15.81) |
| O2 SATURATION VENOUS | 79.36 (22.51) | 79.4 (22.81) |
| O2SAT_MIN | 81.2 (20.03) | 81.09 (20.3) |
| O2_SAT | 93.06 (7.6) | 92.79 (7.82) |
| PC02 VENOUS | 49.31 (15.62) | 49.0 (15.8) |
| PH VENOUS | 7.42 (0.08) | 7.42 (0.08) |

| | | |
|---|---|---|
| PLATELET | 180.5 (88.63) | 183.73 (90.44) |
| PO2 VENOUS | 78.62 (55.71) | 79.92 (56.74) |
| POTASSIUM | 5.31 (1.62) | 5.3 (1.77) |
| PROCALCITONIN | 7.0 (36.47) | 6.72 (35.22) |
| PTT | 53.95 (38.33) | 53.19 (38.06) |
| RBC COUNT | 4.5 (0.77) | 4.54 (0.75) |
| RESPIRATORY_RATE | 21.34 (5.89) | 21.28 (5.77) |
| RESP_RATE_MAX | 33.91 (20.6) | 33.64 (20.02) |
| SERUM CREATININE | 3.21 (3.86) | 3.04 (3.61) |
| SODIUM | 142.84 (8.85) | 142.8 (8.86) |
| SYSTOLIC_BP | 131.03 (25.6) | 130.59 (24.99) |
| SYSTOLIC_BP_MIN | 89.03 (25.38) | 89.76 (25.36) |
| SYSTOL_BP_MAX | 166.41 (26.8) | 165.08 (26.97) |
| TEMPERATURE | 98.98 (1.97) | 99.06 (2.11) |
| TEMP_MAX | 100.86 (2.08) | 100.93 (2.17) |
| TOTAL BILIRUBIN | 1.25 (2.64) | 1.26 (2.95) |
| TOTAL_PROTEIN | 7.2 (0.9) | 7.23 (0.91) |
| WBC | 14.82 (10.73) | 14.91 (10.85) |

#### Categorical Variables

| | | |
|---|---|---|
| **ACUTE KIDNEY INJURY, No (%)** | | |
| No | 3128 (92.5) | 2320 (92.4) |
| Yes | 254 (7.5) | 190 (7.6) |
| **ACUTE MI, No (%)** | | |
| No | 3316 (98.0) | 2482 (98.9) |
| Yes | 66 ( 2.0) | 28 (1.1) |
| **ACUTE VENOUS THROMBOEMBOLISM, No (%)** | | |
| No | 3354 ( 99.2) | 2488 (99.1) |
| Yes | 28 ( 0.8) | 22 (0.9) |
| **ALCOHOLIC NONALCOHOLIC LIVER DISEASE, No (%)** | | |
| No | 3308 ( 97.8) | 2451 (97.6) |
| Yes | 74 ( 2.2) | 59 (2.4) |
| **ARDS, No (%)** | | |
| No | 3358 ( 99.3) | 2492 (99.3) |
| Yes | 24 ( 0.7) | 18 (0.7) |
| **ASTHMA, No (%)** | | |
| No | 3225 ( 95.4) | 2418 (96.3) |
| Yes | 157 ( 4.6) | 92 (3.7) |
| **CANCER FLAG, No (%)** | | |
| No | 3073 ( 90.9) | 2304 (91.8) |
| Yes | 309 ( 9.1) | 206 (8.2) |
| **CEREBRAL INFARCTION, No (%)** | | |
| No | 3346 ( 98.9) | 2486 (99.0) |
| Yes | 36 ( 1.1) | 24 (1.0) |
| **CHRONIC KIDNEY DISEASE, No (%)** | | |

| | | |
|---|---|---|
| No | 2898 ( 85.7) | 2277 (90.7) |
| Yes | 484 ( 14.3) | 233 (9.3) |
| **CHRONIC VIRAL HEPATITIS, No (%)** | | |
| No | 3346 ( 98.9) | 2483 (98.9) |
| Yes | 36 ( 1.1) | 27 (1.1) |
| **COPD, No (%)** | | |
| No | 3205 ( 94.8) | 2434 (97.0) |
| Yes | 177 ( 5.2) | 76 (3.0) |
| **CROHNS DISEASE, No (%)** | | |
| No | 3375 ( 99.8) | 2505 (99.8) |
| Yes | 7 ( 0.2) | 5 (0.2) |
| **DIABETES, No (%)** | | |
| No | 2484 ( 73.4) | 1998 (79.6) |
| Yes | 898 ( 26.6) | 512 (20.4) |
| **ENCOUNTER TYPE, No (%)** | | |
| Inpatient | 3147 (93.1) | 2337 (93.1) |
| Other | 235 (6.9) | 173 (6.9) |
| **HIV FLAG, No (%)** | | |
| No | 3317 ( 98.1) | 2461 (98.0) |
| Yes | 65 ( 1.9) | 49 (2.0) |
| **HYPERTENSION, No (%)** | | |
| No | 1991 ( 58.9) | 1734 (69.1) |
| Yes | 1391 ( 41.1) | 776 (30.9) |
| **ICU, No (%)** | | |
| No | 2429 ( 71.8) | 1813 (72.2) |
| Yes | 707 ( 20.9) | 514 (20.5) |
| **INPATIENT NON ICU, No (%)** | | |
| No | 573 (16.9) | 453 (18.0) |
| Yes | 2563 ( 75.8) | 1874 (74.7) |
| **INTRACEREBRAL HEMORRHAGE, No (%)** | | |
| No | 3378 ( 99.9) | 2506 (99.8) |
| Yes | 4 (0.1) | 4 (0.2) |
| **OBESITY, No (%)** | | |
| No | 3100 ( 91.7) | 2351 (93.7) |
| Yes | 282 ( 8.3) | 159 (6.3) |
| **OBSTRUCTIVE SLEEP APNEA, No (%)** | | |
| No | 3302 ( 97.6) | 2469 (98.4) |
| Yes | 80 ( 2.4) | 41 (1.6) |
| **RACE ETHNICITY COMBINED, No (%)** | | |
| ASIAN | 157 (4.6) | 118 (4.7) |
| BLACK OR AFRICAN-AMERICAN | 918 (27.1) | 694 (27.6) |
| HISPANIC | 923 (27.3) | 686 (27.3) |
| WHITE | 798 (23.6) | 538 (21.4) |
| OTHER | 457 (13.5) | 367 (14.6) |
| NATIVE HAWAIIAN OR PACIFIC ISLANDER | 1 (0.0) | 1 (0.0) |

| | | |
|---|---|---|
| AMERICAN INDIAN OR ALASKA NATIVE | 1 (0.0) | 1 (0.0) |
| **SEX, No (%)** | | |
| MALE | 1993 ( 58.9) | 1475 (58.8) |
| FEMALE | 1389 ( 41.1) | 1035 (41.2) |
| **SMOKING , No (%)** | | |
| NEVER | 1701 ( 50.3) | 1266 (50.4) |
| Current | 153 (4.5) | 117 (4.7) |
| Past | 807 (23.9) | 483 (19.2) |
| PASSIVE | 2 (0.1) | 1 (0.0) |
| **ULCERATIVE COLITIS, No (%)** | | |
| No | 3369 ( 99.6) | 2501 (99.6) |
| Yes | 13 ( 0.4) | 9 (0.4) |

*P-value from student t-test for continuous variables and chisquare test for categorical variables

| data | | | Test Set (n = 846), 20% of 15th July data | |
|---|---|---|---|---|
| **With Cardiac Disease (n =872 )** | **P-values*** | | **Total (n=846)** | **Without Cardiac Disease (n = 653)** |
| 73.27 (12.55) | 3.04E-35 | | 67.47 (15.71) | 65.88 (16.38) |
| 2.46 (0.67) | 0.720983 | | 2.44 (0.69) | 2.45 (0.69) |
| 128.9 (421.48) | 0.134413 | | 128.37 (517.97) | 119.3 (540.1) |
| 17.12 (5.29) | 0.154926 | | 16.79 (5.1) | 16.52 (5.09) |
| 224.71 (853.44) | 0.223857 | | 224.13 (1291.02) | 218.91 (1427.38) |
| 0.81 (0.93) | 0.160497 | | 0.87 (2.63) | 0.88 (2.95) |
| 28.54 (7.38) | 0.761675 | | 28.83 (7.46) | 29.32 (7.67) |
| 985.62 (1903.89) | 8.21E-19 | | 513.83 (1279.54) | 294.63 (764.67) |
| 62.47 (43.54) | 4.03E-07 | | 55.54 (44.87) | 53.05 (45.01) |
| 174.42 (116.42) | 7.79E-05 | | 195.09 (127.66) | 204.24 (130.53) |
| 8.93 (0.88) | 0.697778 | | 8.96 (0.82) | 8.95 (0.83) |
| 109.67 (8.88) | 0.252780 | | 110.11 (8.65) | 110.08 (8.58) |
| 5.0 (5.05) | 0.005945 | | 5.59 (5.45) | 5.93 (5.62) |
| 73.34 (16.15) | 0.229716 | | 73.54 (15.2) | 74.06 (14.49) |
| 46.16 (15.28) | 2.03E-06 | | 48.33 (15.59) | 49.14 (15.85) |
| 98.73 (17.85) | 0.000283 | | 96.98 (17.73) | 96.87 (17.51) |
| 30.28 (23.63) | 3.91E-11 | | 35.41 (28.73) | 37.87 (29.86) |
| 0.2 (0.31) | 0.759916 | | 0.22 (0.49) | 0.23 (0.53) |
| 2.52 (3.46) | 0.014025 | | 2.56 (3.53) | 2.47 (3.45) |
| 1053.59 (1716.18) | 0.126327 | | 1236.76 (2344.07) | 1039.09 (1330.44) |
| 280.74 (188.93) | 0.281756 | | 274.94 (195.55) | 271.99 (197.48) |
| 27.19 (5.81) | 0.000601 | | 26.38 (5.33) | 26.35 (5.19) |
| 91.56 (21.77) | 2.54E-10 | | 97.18 (20.33) | 98.92 (20.08) |
| 121.3 (28.3) | 0.189528 | | 120.38 (24.43) | 120.44 (23.97) |
| 10.14 (2.72) | 1.11E-07 | | 10.54 (2.78) | 10.67 (2.8) |
| 1.8 (1.58) | 1.30E-06 | | 1.51 (0.96) | 1.44 (0.92) |
| 318.14 (839.28) | 0.066442 | | 373.43 (994.97) | 385.3 (1073.65) |
| 689.58 (734.37) | 0.037313 | | 716.0 (927.29) | 708.12 (960.19) |
| 1.64 (1.78) | 0.227937 | | 1.7 (1.35) | 1.7 (1.36) |
| 20.83 (11.49) | 0.495127 | | 21.65 (12.33) | 21.71 (12.25) |
| 30.59 (2.97) | 0.910013 | | 30.36 (2.79) | 30.31 (2.75) |
| 33.45 (1.41) | 0.000131 | | 33.62 (1.4) | 33.7 (1.37) |
| 93.76 (8.1) | 0.018952 | | 92.36 (7.5) | 91.96 (7.34) |
| 9.83 (1.64) | 7.49E-06 | | 9.58 (1.6) | 9.5 (1.61) |
| 0.9 (0.53) | 0.473918 | | 0.9 (0.55) | 0.91 (0.54) |
| 10.62 (5.53) | 0.003678 | | 10.15 (4.75) | 10.08 (4.73) |
| 18.02 (22.19) | 0.063645 | | 18.81 (22.93) | 19.11 (23.17) |
| 66.06 (15.73) | 0.432086 | | 65.89 (15.73) | 65.89 (16.1) |
| 79.26 (21.65) | 0.882016 | | 79.93 (22.03) | 79.72 (21.94) |
| 81.55 (19.23) | 0.561668 | | 82.18 (18.58) | 82.48 (18.27) |
| 93.82 (6.86) | 0.000385 | | 93.37 (7.18) | 92.91 (7.65) |
| 50.17 (15.08) | 0.078335 | | 48.71 (14.47) | 48.34 (14.34) |
| 7.42 (0.08) | 0.378739 | | 7.42 (0.07) | 7.43 (0.07) |

| | | | | |
|---|---|---|---|---|
| 171.23 (82.58) | 0.000181 | | 181.56 (85.45) | 186.18 (85.81) |
| 75.08 (52.69) | 0.045458 | | 78.07 (55.87) | 75.94 (54.1) |
| 5.36 (1.05) | 0.238833 | | 5.28 (1.06) | 5.24 (1.04) |
| 7.83 (40.02) | 0.518561 | | 9.54 (43.63) | 10.24 (46.62) |
| 56.09 (39.04) | 0.098288 | | 53.48 (36.92) | 51.81 (35.49) |
| 4.39 (0.83) | 5.97E-06 | | 4.57 (0.78) | 4.62 (0.74) |
| 21.52 (6.22) | 0.347297 | | 21.61 (5.83) | 21.8 (5.98) |
| 34.7 (22.2) | 0.227068 | | 33.53 (19.68) | 33.58 (18.94) |
| 3.7 (4.48) | 8.19E-05 | | 3.2 (3.6) | 2.94 (3.41) |
| 142.95 (8.84) | 0.681707 | | 142.73 (8.23) | 142.68 (8.28) |
| 132.33 (27.27) | 0.109561 | | 130.83 (26.08) | 130.65 (25.09) |
| 86.92 (25.31) | 0.005323 | | 89.25 (24.49) | 90.7 (24.88) |
| 170.24 (25.94) | 1.07E-06 | | 165.42 (26.68) | 165.22 (26.46) |
| 98.74 (1.48) | 1.35E-06 | | 99.04 (2.78) | 99.2 (1.76) |
| 100.66 (1.8) | 0.000517 | | 100.93 (2.09) | 101.03 (2.19) |
| 1.22 (1.43) | 0.653973 | | 1.17 (1.59) | 1.15 (1.73) |
| 7.11 (0.88) | 0.001205 | | 7.26 (0.92) | 7.24 (0.87) |
| 14.54 (10.37) | 0.362529 | | 14.66 (9.44) | 14.8 (9.68) |

| | | | | |
|---|---|---|---|---|
| | 0.882585941 | | | |
| (808 ( 92.7) | | | 789 ( 93.3) | 611 ( 93.6) |
| 64 ( 7.3 ) | | | 57 ( 6.7) | 42 ( 6.4) |
| | 5.86E-09 | | | |
| (834 ( 95.6) | | | 832 ( 98.3) | 645 ( 98.8) |
| 38 ( 4.4 ) | | | 14 ( 1.7) | 8 ( 1.2) |
| | 0.754975725 | | | |
| (866 ( 99.3) | | | 839 ( 99.2) | 646 ( 98.9) |
| 6 ( 0.7 ) | | | 7 ( 0.8) | 7 ( 1.1) |
| | 0.336100166 | | | |
| (857 ( 98.3) | | | 812 ( 96.0) | 631 ( 96.6) |
| 15 ( 1.7 ) | | | 34 ( 4.0) | 22 ( 3.4) |
| | 0.883856262 | | | |
| (866 ( 99.3) | | | 837 ( 98.9) | 645 ( 98.8) |
| 6 ( 0.7 ) | | | 9 ( 1.1) | 8 ( 1.2) |
| | 7.20E-06 | | | |
| (807 ( 92.5) | | | 798 ( 94.3) | 624 ( 95.6) |
| 65 ( 7.5 ) | | | 48 ( 5.7) | 29 ( 4.4) |
| | 0.001842591 | | | |
| (769 ( 88.2) | | | 774 ( 91.5) | 607 ( 93.0) |
| 103 ( 11.8 ) | | | 72 ( 8.5) | 46 ( 7.0) |
| | 0.395566942 | | | |
| (860 ( 98.6) | | | 837 ( 98.9) | 645 ( 98.8) |
| 12 ( 1.4 ) | | | 9 ( 1.1) | 8 ( 1.2) |
| | 3.25E-45 | | | |

| | | | | |
|---|---|---|---|---|
| (621 ( 71.2) | | | 706 ( 83.5) | 587 ( 89.9) |
| 251 ( 28.8 ) | | | 140 ( 16.5) | 66 ( 10.1) |
| | 0.933475607 | | | |
| (863 ( 99.0) | | | 837 ( 98.9) | 647 ( 99.1) |
| 9 ( 1.0 ) | | | 9 ( 1.1) | 6 ( 0.9) |
| | 3.53E-22 | | | |
| (771 ( 88.4) | | | 817 ( 96.6) | 638 ( 97.7) |
| 101 ( 11.6 ) | | | 29 ( 3.4) | 15 ( 2.3) |
| | 0.792032466 | | | |
| (870 ( 99.8) | | | 844 ( 99.8) | 653 ( 100.0) |
| 2 ( 0.2 ) | | | 2 ( 0.2) | 0 (0.0) |
| | 9.52E-43 | | | |
| (486 ( 55.7) | | | 626 ( 74.0) | 521 ( 79.8) |
| 386 ( 44.3 ) | | | 220 ( 26.0) | 132 ( 20.2) |
| | 0.888291625 | | | |
| 810 (92.9) | | | 789 (93.3) | 610 (93.4) |
| 62 (7.1) | | | 57 (6.7) | 43 (6.6) |
| | 0.940816165 | | | |
| 856 (98.2) | | | 837 (98.9) | 645 ( 98.8) |
| 16 (1.8) | | | 9 (1.1) | 8 ( 1.2) |
| | 7.58E-93 | | | |
| 257 (29.5) | | | 505 (59.7) | 446 ( 68.3) |
| 615 (70.5) | | | 341 (40.3) | 207 ( 31.7) |
| | 0.323233683 | | | |
| 616 (70.6) | | | 598 (70.7) | 460 ( 70.4) |
| 193 (22.1) | | | 190 (22.5) | 149 ( 22.8) |
| | 0.003910682 | | | |
| 120 (13.8) | | | 144 (17.0) | 116 (17.8) |
| 689 (79.0) | | | 644 (76.1) | 493 (75.5) |
| | 0.543395076 | | | |
| 872 (100.0) | | | 844 (99.8) | 652 (99.8) |
| 0 (0.0) | | | 2 (0.2) | 1 (0.2) |
| | 1.45E-12 | | | |
| 749 (85.9) | | | 772 (91.3) | 594 (91.0) |
| 123 (14.1) | | | 74 (8.7) | 59 (9.0) |
| | 3.78E-06 | | | |
| 833 (95.5) | | | 827 (97.8) | 640 (98.0) |
| 39 (4.5) | | | 19 (2.2) | 13 (2.0) |
| | 5.13E-05 | | | |
| 39 (4.5) | | | 47 (5.6) | 34 (5.2) |
| 224 (25.7) | | | 221 (26.1) | 178 (27.3) |
| 237 (27.2) | | | 236 (27.9) | 196 (30.0) |
| 260 (29.8) | | | 169 (20.0) | 108 (16.5) |
| 90 (10.3) | | | 139 (16.4) | 114 (17.5) |
| 0 (0.0) | | | 0 (0.0) | 0 (0.0) |

| | | | | |
|---|---|---|---|---|
| 0 (0.0) | | | 0 (0.0) | 0 (0.0) |
| | 0.77155788 | | | |
| 518 (59.4) | | | 497 (58.7) | 386 (59.1) |
| 354 (40.6) | | | 349 (41.3) | 267 (40.9) |
| | 9.65E-13 | | | |
| 435 (49.9) | | | 439 (51.9) | 345 (52.8) |
| 36 (4.1) | | | 37 (4.4) | 26 (4.0) |
| 324 (37.2) | | | 187 (22.1) | 121 (18.5) |
| 1 (0.1) | | | 0 (0.0) | 0 (0.0) |
| | 0.925026688 | | | |
| 868 (99.5) | | | 846 (100.0) | 653 (100.0) |
| 4 (0.5) | | | 0 (0.0) | 0 (0.0) |

| | |
|---|---|
| **With Cardiac Disease (n = 193)** | **P-values*** |
| 72.85 (11.76) | 1.44E-10 |
| 2.42 (0.69) | 0.616719 |
| 158.17 (437.36) | 0.310716 |
| 17.7 (5.04) | 0.004923 |
| 241.51 (661.54) | 0.763038 |
| 0.81 (0.93) | 0.565992 |
| 27.4 (6.63) | 0.001363 |
| 1120.09 (2016.66) | 1.29E-06 |
| 63.96 (43.43) | 0.002619 |
| 163.3 (111.79) | 9.26E-05 |
| 9.0 (0.79) | 0.424453 |
| 110.19 (8.87) | 0.876153 |
| 4.35 (4.57) | 0.000498 |
| 71.77 (17.35) | 0.108707 |
| 45.6 (14.35) | 0.004165 |
| 97.37 (18.51) | 0.743188 |
| 27.78 (23.41) | 5.99E-06 |
| 0.22 (0.3) | 0.798222 |
| 2.87 (3.81) | 0.191102 |
| 1915.9 (4216.83) | 0.009662 |
| 284.9 (189.05) | 0.411366 |
| 26.47 (5.8) | 0.811889 |
| 91.31 (20.15) | 1.01E-05 |
| 120.19 (25.98) | 0.906720 |
| 10.1 (2.71) | 0.010942 |
| 1.74 (1.04) | 0.001209 |
| 330.66 (637.93) | 0.530598 |
| 743.53 (804.15) | 0.647665 |
| 1.67 (1.34) | 0.808482 |
| 21.43 (12.65) | 0.787784 |
| 30.5 (2.92) | 0.430252 |
| 33.34 (1.44) | 0.002191 |
| 93.71 (7.87) | 0.006400 |
| 9.84 (1.52) | 0.008070 |
| 0.9 (0.57) | 0.902433 |
| 10.4 (4.84) | 0.420691 |
| 17.8 (22.13) | 0.478494 |
| 65.87 (14.41) | 0.986668 |
| 80.68 (22.38) | 0.642645 |
| 81.2 (19.6) | 0.431577 |
| 94.92 (4.99) | 3.42E-05 |
| 49.98 (14.85) | 0.221491 |
| 7.42 (0.07) | 0.165882 |

| | |
|---|---|
| 166.03 (82.55) | 0.003407 |
| 85.32 (61.15) | 0.096001 |
| 5.43 (1.11) | 0.036127 |
| 7.11 (31.06) | 0.331820 |
| 58.96 (40.93) | 0.058061 |
| 4.39 (0.86) | 0.000913 |
| 20.98 (5.25) | 0.075160 |
| 33.38 (22.06) | 0.911150 |
| 4.07 (4.07) | 0.000527 |
| 142.9 (8.07) | 0.754595 |
| 131.48 (29.28) | 0.729811 |
| 84.36 (22.53) | 0.001073 |
| 166.11 (27.44) | 0.693991 |
| 98.47 (4.8) | 0.044077 |
| 100.61 (1.72) | 0.006975 |
| 1.22 (1.02) | 0.486184 |
| 7.3 (1.09) | 0.538424 |
| 14.2 (8.59) | 0.407910 |

| | |
|---|---|
| | 0.62476289 |
| 178 ( 92.2) | |
| 15 ( 7.8) | |
| | 0.13858252 |
| 187 ( 96.9) | |
| 6 ( 3.1) | |
| | 0.32113486 |
| 193 ( 100.0) | |
| 0 (0.0) | |
| | 0.11837316 |
| 181 ( 93.8) | |
| 12 ( 6.2) | |
| | 0.65864418 |
| 192 ( 99.5) | |
| 1 ( 0.5) | |
| | 0.00750029 |
| 174 ( 90.2) | |
| 19 ( 9.8) | |
| | 0.00771183 |
| 167 ( 86.5) | |
| 26 ( 13.5) | |
| | 0.65864418 |
| 192 ( 99.5) | |
| 1 ( 0.5) | |
| | 5.04E-20 |

| | |
|---|---|
| 119 ( 61.7) | |
| 74 ( 38.3) | |
| | 0.7212213 |
| 190 ( 98.4) | |
| 3 ( 1.6) | |
| | 0.00193523 |
| 179 ( 92.7) | |
| 14 ( 7.3) | |
| | 0.07826238 |
| 191 ( 99.0) | |
| 2 ( 1.0) | |
| | 3.20E-12 |
| 105 ( 54.4) | |
| [88 ( 45.6) | |
| | 0.87109763 |
| 179 (92.7) | |
| 14 (7.3) | |
| | 0.65864418 |
| 192 ( 99.5) | |
| 1 (0.5) | |
| | 1.34E-20 |
| 59 (30.6) | |
| 134 (69.4) | |
| | 0.74146061 |
| 138 ( 71.5) | |
| 41 (21.2) | |
| | 0.35425853 |
| 28 (14.5) | |
| 151 (78.2) | |
| | 0.94118182 |
| 192 ( 99.5) | |
| 1 (0.5) | |
| | 0.68862702 |
| 178 ( 92.2) | |
| 15 (7.8) | |
| | 0.51927591 |
| 187 ( 96.9) | |
| 6 (3.1) | |
| | 3.70E-05 |
| 13 (6.7) | |
| 43 (22.3) | |
| 40 (20.7) | |
| 61 (31.6) | |
| 25 (13.0) | |
| 0 (0.0) | |

| | |
|---|---|
| 0 (0.0) | |
| | 0.75413949 |
| 111 ( 57.5) | |
| 82 (42.5) | |
| | 0.00115863 |
| 94, 48.7 | |
| 11, 5.7 | |
| 66, 34.2 | |
| 0 (0.0) | |
| 193 (100.0) | 1 |
| 0 (0.0) | |

| TABLE S2- Variables (Continous Variables) | Development Set (n = 3382) | Test Set (n = 846) |
|---|---|---|
| AGE | 0 | 0 |
| ALBUMIN | 86 | 27 |
| ALT | 92 | 27 |
| ANION_GAP | 8 | 6 |
| AST | 137 | 37 |
| BASOPHIL (%) | 35 | 22 |
| BMI | 520 | 136 |
| BRAIN NATRIURETIC PROTEIN | 980 | 251 |
| BUN | 7 | 5 |
| C-REACTIVE PROTEIN | 468 | 116 |
| CALCIUM | 5 | 5 |
| CHLORIDE | 4 | 3 |
| D-DIMER | 661 | 162 |
| DIASTOLIC_BP | 230 | 53 |
| DIASTOLIC_BP_MIN | 142 | 30 |
| DIASTOL_BP_MAX | 142 | 30 |
| EGFR | 565 | 137 |
| EOSINOPHIL # | 79 | 22 |
| EOSINOPHIL (%) | 35 | 15 |
| FERRITIN | 499 | 123 |
| GLUCOSE | 4 | 5 |
| HCO3 VENOUS | 772 | 173 |
| HEART_RATE | 209 | 46 |
| HEART_RATE_MAX | 143 | 30 |
| HEMOGLOBIN | 7 | 4 |
| INR | 832 | 206 |
| INTERLEUKIN-6 | 1589 | 404 |
| LDH | 617 | 163 |
| LYMPHOCYTE # | 79 | 22 |
| LYMPHOCYTE (%) | 29 | 13 |
| MCH | 7 | 4 |
| MCHC | 7 | 4 |
| MCV | 7 | 4 |
| MEAN PLATELET VOLUME (MPV) | 21 | 9 |
| MONOCYTE # | 79 | 22 |
| MONOCYTE (%) | 29 | 13 |
| NEUTROPHIL # | 11 | 9 |
| NEUTROPHIL (%) | 29 | 13 |
| O2 SATURATION VENOUS | 794 | 189 |
| O2SAT_MIN | 211 | 46 |
| O2_SAT | 211 | 46 |
| PC02 VENOUS | 671 | 158 |
| PH VENOUS | 671 | 158 |
| PLATELET | 11 | 4 |

| | | |
|---|---|---|
| PO2 VENOUS | 852 | 204 |
| POTASSIUM | 17 | 5 |
| PROCALCITONIN | 642 | 160 |
| PTT | 866 | 219 |
| RBC COUNT | 7 | 4 |
| RESPIRATORY_RATE | 220 | 50 |
| RESP_RATE_MAX | 146 | 33 |
| SERUM CREATININE | 7 | 5 |
| SODIUM | 427 | 95 |
| SYSTOLIC_BP | 230 | 53 |
| SYSTOLIC_BP_MIN | 142 | 30 |
| SYSTOL_BP_MAX | 142 | 30 |
| TEMPERATURE | 209 | 46 |
| TEMP_MAX | 209 | 46 |
| TOTAL BILIRUBIN | 86 | 29 |
| TOTAL_PROTEIN | 104 | 32 |
| WBC | 8 | 4 |

#### Categorical Variables

| | | |
|---|---|---|
| ACUTE KIDNEY INJURY | 0 | 0 |
| ACUTE MI | 0 | 0 |
| ACUTE VENOUS THROMBOEMBOLISM | 0 | 0 |
| ALCOHOLIC NONALCOHOLIC LIVER DISEASE | 0 | 0 |
| ARDS | 0 | 0 |
| ASTHMA | 0 | 0 |
| CANCER FLAG | 0 | 0 |
| CEREBRAL INFARCTION | 0 | 0 |
| CHRONIC KIDNEY DISEASE | 0 | 0 |
| CHRONIC VIRAL HEPATITIS | 0 | 0 |
| COPD | 0 | 0 |
| CROHNS DISEASE | 0 | 0 |
| DIABETES | 0 | 0 |
| ENCOUNTER TYPE | 0 | 0 |
| HIV FLAG | 0 | 0 |
| HYPERTENSION | 0 | 0 |
| ICU | 246 | 58 |
| INPATIENT NON ICU | 246 | 58 |
| INTRACEREBRAL HEMORRHAGE | 0 | 0 |
| OBESITY | 0 | 0 |
| OBSTRUCTIVE SLEEP APNEA | 0 | 0 |
| RACE ETHNICITY COMBINED | 127 | 34 |
| SEX | 0 | 0 |
| SMOKING | 719 | 183 |
| ULCERATIVE COLITIS | 0 | 0 |
